# Generalisable functional imaging classifiers of schizophrenia have multifunctionality as trait, state, and staging biomarkers

**DOI:** 10.1101/2024.01.02.23300101

**Authors:** Takahiko Kawashima, Ayumu Yamashita, Yujiro Yoshihara, Yuko Kobayashi, Naohiro Okada, Kiyoto Kasai, Ming-Chyi Huang, Akira Sawa, Junichiro Yoshimoto, Okito Yamashita, Toshiya Murai, Jun Miyata, Mitsuo Kawato, Hidehiko Takahashi

## Abstract

Schizophrenia spectrum disorder (SSD) is one of the top causes of disease burden; similar to other psychiatric disorders, SSD lacks widely applicable and objective biomarkers. This study aimed to introduce a novel resting-state functional connectivity (rs-FC) magnetic resonance imaging (MRI) biomarker for diagnosing SSD. It was developed using customised machine learning on an anterogradely and retrogradely harmonised dataset from multiple sites, including 617 healthy controls and 116 patients with SSD. Unlike previous rs-FC MRI biomarkers, this new biomarker demonstrated a notable accuracy rate of 77.3% in an independent validation cohort, including 404 healthy controls and 198 patients with SSD from seven different sites, effectively mitigating across-scan variability. Importantly, our biomarker specifically identified SSD, differentiating it from other psychiatric disorders. Our analysis identified 47 important FCs significant in SSD classification, several of which are involved in SSD pathophysiology. Beyond their potential as trait markers, we explored the utility of these FCs as both state and staging markers. First, based on aggregated FCs, we built prediction models for clinical scales of trait and/or state. Thus, we successfully predicted delusional inventory scores (*r*=0.331, *P*=0.0177), but not the overall symptom severity (*r*=0.128, *P*=0.178). Second, through comprehensive analysis, we uncovered associations between individual FCs and symptom scale scores or disease stages, presenting promising candidate FCs for state or staging markers. This study underscores the potential of rs-FC as a clinically applicable neural phenotype marker for SSD and provides actionable targets to neuromodulation therapies.

## INTRODUCTION

Diagnosis of schizophrenia spectrum disorder (SSD) relies on diagnostic criteria that categorise disorders based solely on signs and symptoms. There are currently no widely applicable objective biomarkers^1^ that can distinguish individuals with SSD from their healthy counterparts.

Generating an SSD classifier using resting-state functional connectivity (rs-FC) magnetic resonance imaging (MRI) is an emerging research topic. However, the practical use of these classifiers is hindered by several key challenges^2–5^. First, there is an accuracy problem. In studies with a large sample size and external validation^6,7^, classifier performance was below biomarker requirements (approximately 80%). Accuracy typically decreases with increase in sample size (*N*>200) in whole-brain imaging biomarkers^8^. A machine learning algorithm requiring many explanatory variables (i.e. FCs) needs more data to achieve accuracy and generalisability; however, large training samples from multiple facilities may cause a large site effect, deteriorating data quality^9^. Second, there is an issue with generalisability. Most previous studies have used only internal or limited external validation and lacked genuinely independent cohorts obtained in multiple sites not involved in the initial research^10^. Third, across-session variability poses a problem. The reliability of rs-fMRI is questionable over repeated tests^11^. Finally, there is a lack of specificity in potential biomarkers identified in rs-FC MRI studies focusing on specific disorders. Overcoming these issues is crucial for determining truly effective biomarkers for clinical use in SSD.

Although disease trait markers assist early intervention, state markers estimate dynamic changes in response to treatment^12,13^ and staging markers aid disease prevention or personalising interventions^14^. In SSD research, few studies have succeeded in developing state markers using neuroimaging, and for staging markers, there is no consensus on biological staging models^15^.

Patient biological data may simultaneously encompass information on traits, states, and disease stages. As machine learning exploits these features without making distinctions, diagnostic markers developed using machine learning may also function as state and/or staging markers. Previous rs-FC biomarker studies reported multiple aggregated FCs as biomarkers^16,17^; however, whether individual FCs can be used as one or more of these three types of biomarkers remains unexplored. A previous study showed that FCs originally identified as diagnostic markers dynamically changed after treatment^18^; thus, they also functioned as state markers. Nevertheless, thoroughly investigating individual FCs has not been attempted, making it difficult to determine whether any FCs can benefit early disease detection and intervention as a trait marker or treatment target selection and drug discovery as a surrogate marker.

Our study focused on two key areas (**Figure 1**). First, we aimed to develop a clinically viable rs-FC biomarker for SSD, addressing the challenges mentioned above. Second, we aimed to investigate whether the diagnostic biomarker can be used as state and staging markers. The findings of this study can help develop new approaches for improving the diagnosis and treatment of various psychiatric disorders, not only SSD.

**Figure 1.**
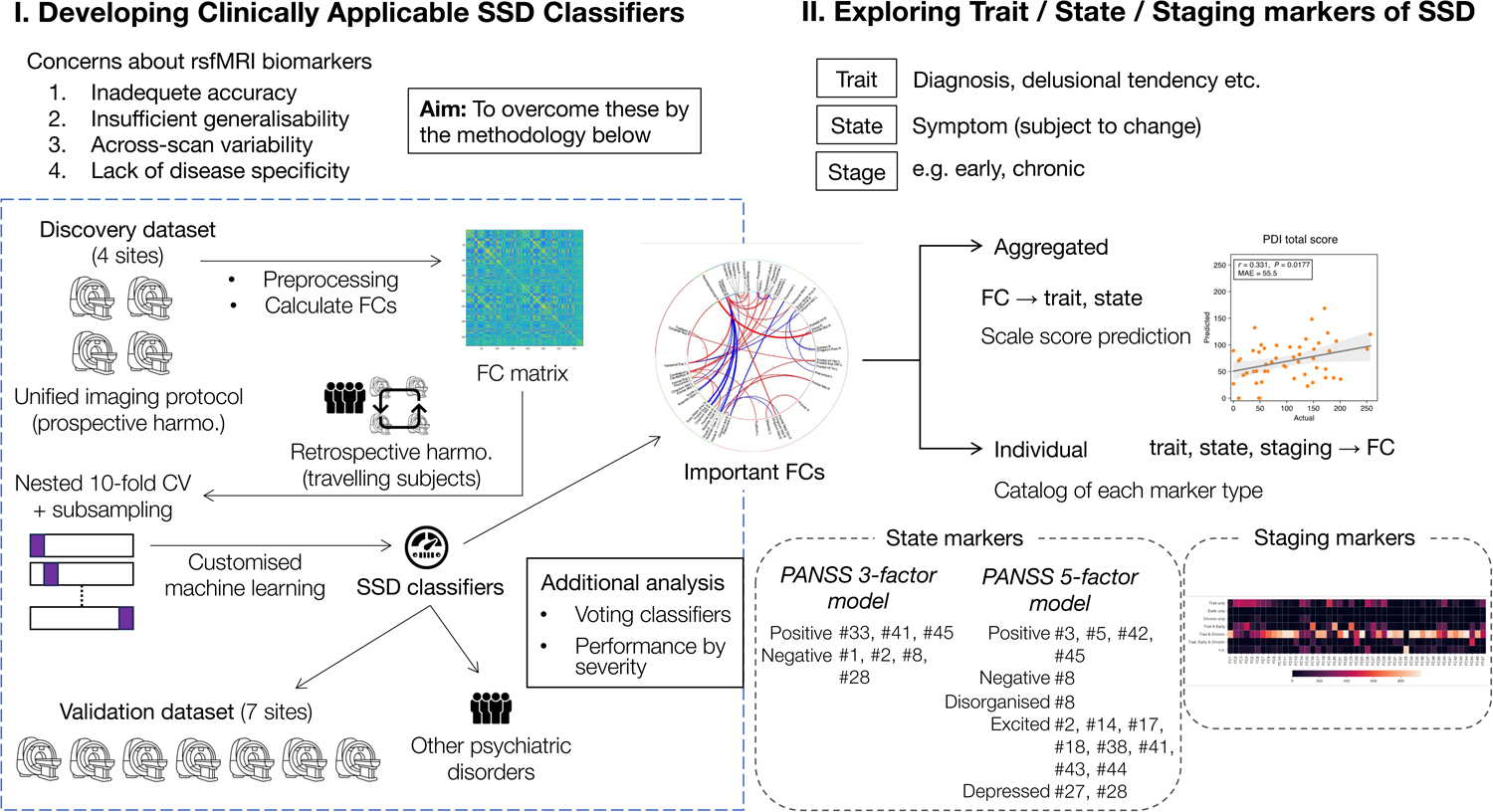
Outline of the study. This study was composed of two parts. (I) Constructing SSD classifier: Using the discovery dataset, we processed rs-fMRI images into an FC matrix for each participant, which was then inputted into machine learning (LASSO) to build SSD classifiers. We obtained the classification performance through 10-fold CV and examined its external generalisability using the validation dataset. ‘Important FCs’ were those that made the highest contribution to the classification. To assess classifier specificity for SSD, we also applied the classifiers to other mental disorders. We performed further analyses on another machine learning method (voting classifiers) and on classification performance by disease severity. (II) Exploring trait/state/staging markers of SSD: We investigated the different types of biomarkers inherent in important FCs. First, we attempted to predict clinical scale scores using aggregated FCs. Second, we searched for individual FCs associated with the state and/or disease stage. CV, cross-validation; SSD, schizophrenia spectrum disorder; LASSO, least absolute shrinkage and selection operator; rs-fMRI, resting-state functional magnetic resonance imaging; FC, functional connectivity.

## MATERIALS AND METHODS

### Participants

We used two independent multi-disorder datasets: one for developing classifiers (‘discovery dataset’) and the other for validating the classifiers (‘validation dataset’). Participant characteristics are presented in **Table 1** and **Supplementary Table 1**. There was no overlap of participants between the discovery and validation datasets.

**Table 1.**
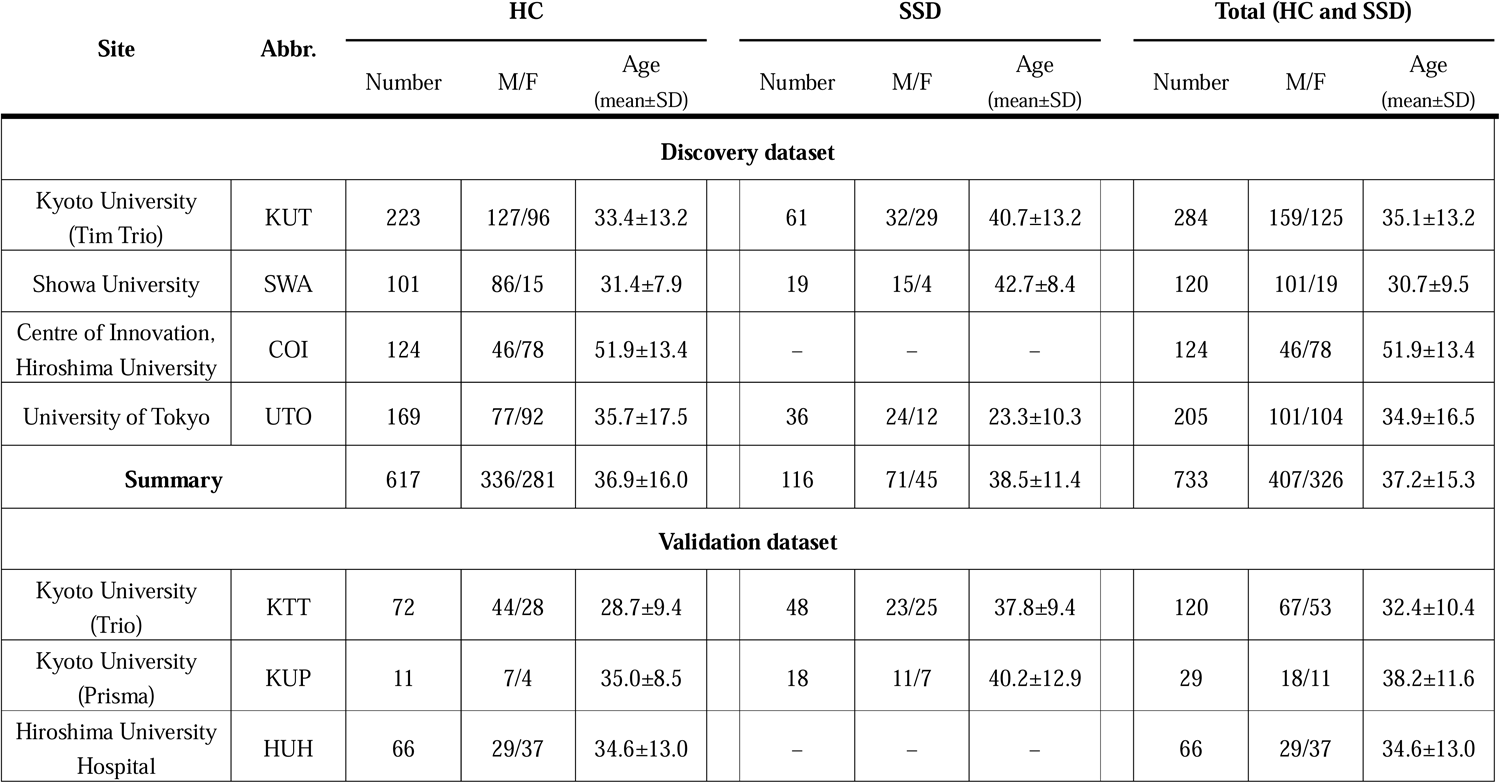
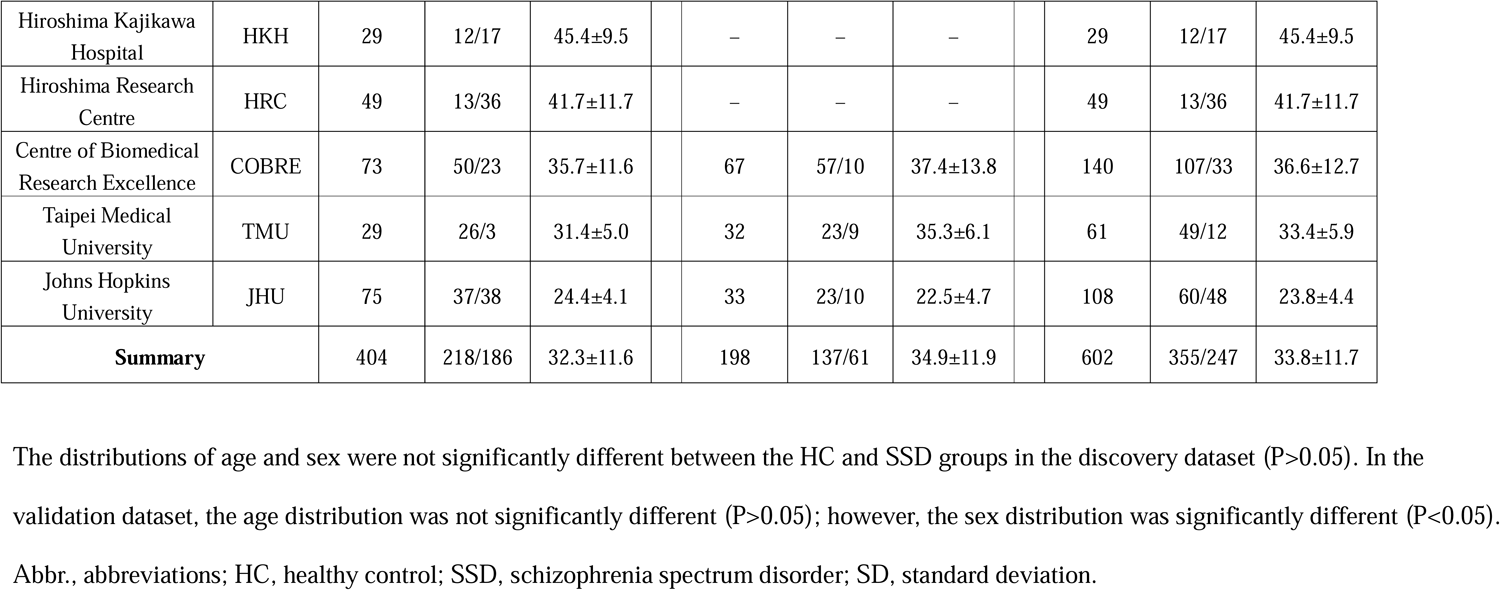
Demographics of the participants (HCs and patients with SSD)

| Site | Abbr. | HC |  |  |  |  | SSD |  |  |  | Total (HC and SSD) |  |  |  |
| --- | --- | --- | --- | --- | --- | --- | --- | --- | --- | --- | --- | --- | --- | --- |
|  |  | Number | M/F | Age<br>(mean±SD) |  |  | Number | M/F | Age<br>(mean±SD) |  |  | Number | M/F | Age<br>(mean±SD) |
| Discovery dataset |  |  |  |  |  |  |  |  |  |  |  |  |  |  |
| Kyoto University<br>(Tim Trio) | KUT | 223 | 127/96 | 33.4±13.2 |  |  | 61 | 32/29 | 40.7±13.2 |  |  | 284 | 159/125 | 35.1±13.2 |
| Showa University | SWA | 101 | 86/15 | 31.4±7.9 |  |  | 19 | 15/4 | 42.7±8.4 |  |  | 120 | 101/19 | 30.7±9.5 |
| Centre of Innovation,<br>Hiroshima University | COI | 124 | 46/78 | 51.9±13.4 |  |  | – | – | – |  |  | 124 | 46/78 | 51.9±13.4 |
| University of Tokyo | UTO | 169 | 77/92 | 35.7±17.5 |  |  | 36 | 24/12 | 23.3±10.3 |  |  | 205 | 101/104 | 34.9±16.5 |
| Summary |  | 617 | 336/281 | 36.9±16.0 |  |  | 116 | 71/45 | 38.5±11.4 |  |  | 733 | 407/326 | 37.2±15.3 |
| Validation dataset |  |  |  |  |  |  |  |  |  |  |  |  |  |  |
| Kyoto University<br>(Trio) | KTT | 72 | 44/28 | 28.7±9.4 |  |  | 48 | 23/25 | 37.8±9.4 |  |  | 120 | 67/53 | 32.4±10.4 |
| Kyoto University<br>(Prisma) | KUP | 11 | 7/4 | 35.0±8.5 |  |  | 18 | 11/7 | 40.2±12.9 |  |  | 29 | 18/11 | 38.2±11.6 |
| Hiroshima University<br>Hospital | HUH | 66 | 29/37 | 34.6±13.0 |  |  | – | – | – |  |  | 66 | 29/37 | 34.6±13.0 |

|  |  |  |  |  |  |  |  |  |  |  |  |  |
| --- | --- | --- | --- | --- | --- | --- | --- | --- | --- | --- | --- | --- |
| Hiroshima Kajikawa Hospital | HKH | 29 | 12/17 | 45.4±9.5 |  | – | – | – |  | 29 | 12/17 | 45.4±9.5 |
| Hiroshima Research Centre | HRC | 49 | 13/36 | 41.7±11.7 |  | – | – | – |  | 49 | 13/36 | 41.7±11.7 |
| Centre of Biomedical Research Excellence | COBRE | 73 | 50/23 | 35.7±11.6 |  | 67 | 57/10 | 37.4±13.8 |  | 140 | 107/33 | 36.6±12.7 |
| Taipei Medical University | TMU | 29 | 26/3 | 31.4±5.0 |  | 32 | 23/9 | 35.3±6.1 |  | 61 | 49/12 | 33.4±5.9 |
| Johns Hopkins University | JHU | 75 | 37/38 | 24.4±4.1 |  | 33 | 23/10 | 22.5±4.7 |  | 108 | 60/48 | 23.8±4.4 |
| **Summary** |  | 404 | 218/186 | 32.3±11.6 |  | 198 | 137/61 | 34.9±11.9 |  | 602 | 355/247 | 33.8±11.7 |

- 2 The distributions of age and sex were not significantly different between the HC and SSD groups in the discovery dataset ( $P>0.05$ ). In the - 3 validation dataset, the age distribution was not significantly different ( $P>0.05$ ); however, the sex distribution was significantly different ( $P<0.05$ ). - 4 Abbr., abbreviations; HC, healthy control; SSD, schizophrenia spectrum disorder; SD, standard deviation.

The discovery dataset comprised data from four sites: Kyoto University Siemens TimTrio scanner, Showa University (SWA), Centre of Innovation in Hiroshima University, and University of Tokyo (UTO). The dataset included 1 045 participants consisting of 617 healthy controls (HCs), 116 patients with SSD (which includes schizophrenia, schizoaffective disorder, and delusional disorder), 148 patients with major depressive disorder (MDD), 125 patients with autism spectrum disorder (ASD), and 39 patients with bipolar disorder (BP).

The validation dataset comprised international cohorts, including the Japanese cohort from Hiroshima Kajikawa Hospital, Hiroshima Rehabilitation Centre, Hiroshima University Hospital, Kyoto University Siemens Trio scanner, and Kyoto University Siemens Prisma scanner. It also included the Taiwanese cohort (Taipei Medical University) and the Centre of Biomedical Research Excellence open dataset (http://fcon_1000.projects.nitrc.org/indi/retro/cobre.html), along with the Johns Hopkins University cohort (United States), which included HCs and patients with early-stage SSD. The total number of participants was 708 (405 HCs, 198 patients with SSD, and 105 patients with MDD).

Clinical scale scores: Positive and Negative Syndrome Scale (PANSS)^19^ and Peters et al. Delusions Inventory (PDI) 21-item version^20^ and information about the dosage of antipsychotics were available from a subset of participants in both datasets (**Supplementary Table 2**).

This study was conducted in accordance with the recommendations of the review boards of institutions affiliated with the principal investigators—namely, Hiroshima University, Kyoto University, SWA, and UTO. Most of the material data in this study can be downloaded from the DecNef Project Brain Data Repository^21^

(https://bicr-resource.atr.jp/srpbsopen/).

### Data acquisition

All data in the discovery dataset were collected using a unified protocol developed by a national project (Strategic Research Program for Brain Science [SRPBS] & Brain/Mind Beyond) in Japan^21^. The MRI data comprised a T1-weighted structural image, rs-fMRI acquired using an echo-planar imaging technique, and field map images. The duration of rs-fMRI was 10 min. The participants were instructed to relax, stay awake, fixate on the central crosshair mark, and not concentrate on specific things. The MRI data for the validation dataset included a structural image and rs-fMRI similar to the discovery dataset; however, some of the data lacked fieldmap images. Scanning parameters for the validation dataset varied by site. The duration of rs-fMRI was approximately 4–6 min. Detailed imaging parameters for both datasets are provided in **Supplementary Table 3**.

### Preprocessing

The data were preprocessed according to a previous report^22^. fMRIPrep version 20.1.11 was used for data preprocessing^23^. First, the first four volumes of the rs-fMRI scan were discarded for T1 equilibration. The preprocessing steps were as follows: slice-timing correction, realignment, coregistration, susceptibility-induced distortion correction using field maps, segmentation of a T1-weighted structural image, normalisation to Montreal Neurological Institute space, and spatial smoothing with an isotropic Gaussian kernel of 6 mm full-width at half-maximum. For the participants without field maps in the validation dataset, we applied ‘fieldmap-less’ distortion correction implemented in fMRIPrep. Further details of the pipeline are available at http://fmriprep.readthedocs.io/en/latest/workflows.html.

### Signal extraction

We used two approaches for fMRI signal extraction: (1) a surface-based approach following the Human Connectome Project pipelines (using the ciftify toolbox version 2.3.2^24^, we converted volume-based MRI data into data based on ‘greyordinate’ [cortical grey matter surface vertices and subcortical grey matter voxels]) and (2) an approach based on the regions of interest (ROIs). For reliable surface-based brain parcellation, we adopted the ROIs from Glasser et al.^25^ (379 parcels in total, comprising 360 cortical parcels as surface ROIs and 19 subcortical parcels as volume ROIs). Using these approaches, we extracted BOLD signal time courses from 379 ROIs. To compare these ROIs with conventional annotations of brain areas and intrinsic brain networks, we referred to the Anatomical Automatic Labelling (AAL) and Ji et al.^26^, respectively.

### Noise removal

We used component-based noise correction (CompCor)^27^ to detect physiological noise. Anatomical CompCor was applied to the subcortical white matter and cerebrospinal fluid regions, and the top five principal components were estimated as physiological noise, except for one participant who had only four components. Accordingly, we regressed out these components together with six head-motion parameters and averaged the signals over the entire brain.

### Temporal filtering

We used temporal bandpass filtering ranging from 0.01 to 0.08 Hz^28^ to the time series of rs-fMRI to extract the low-frequency brain activity characterising resting state.

### Data scrubbing

We removed volumes with considerable head motion based on framewise displacement (FD)^29^. FD was calculated as the sum of the absolute displacements in translation and rotation between two consecutive volumes. We removed volumes with FD >0.5 mm, as proposed in a previous study^29^. In addition, participants whose scrubbed volume ratio exceeded the mean +3 S.D. were excluded.

### Calculation of the FC matrices

We defined FC as the temporal correlation of rs-fMRI BOLD signals between two ROIs. We calculated Pearson’s correlation coefficient of the preprocessed BOLD time series between each pair of ROIs out of Glasser’s 379 ROIs. Fisher’s Z-transformed values of the correlation coefficients constituted an FC matrix for each participant, where the total number of FC was 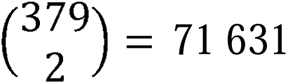.

### Harmonisation of the site differences

We harmonised the site effects in the discovery dataset prospectively^21^ using a unified imaging protocol under the SRPBS & Brain/Mind Beyond project and retrospectively using the travelling subject method^9^. Regarding the travelling subject method, site effects were separately estimated as measurement bias and sampling bias from the rs-fMRI data of these travelling subjects, each of whom underwent scans at multiple sites^9^. We subtracted the estimated measurement bias to obtain harmonised connectivity data (see **Supplementary Text 1**). For the validation dataset, we applied the ComBat harmonisation method^30–32^. In the execution of ComBat harmonisation, we inputted the diagnosis, PANSS total score, PDI total score, age, sex, handedness, and duration of illness (DOI) (for patients with SSD) as auxiliary variables to correct measurement bias.

### Diagnostic classifiers for SSD

Next, we constructed classifiers to differentiate patients with SSD from HCs using machine learning based on 71 631 FC values as features. In subsequent sections, we used only the data of HCs and patients with SSD, except for generalising the models to other disorders. Initially, we used a customised sparse learning algorithm, specifically least absolute shrinkage and selection operator (LASSO), similar to our previous work on MDD^26^. The sparse method in LASSO can prevent overfitting and simultaneously select important features (for the detailed methodology, refer to **Supplementary Text 2**). Although we used LASSO to identify important FCs for SSD classification, we applied a voting method^33^ to enhance the classification performance of the LASSO classifiers.

### Building LASSO classifiers

As illustrated in **Supplementary Figure 1a**, we implemented a nested 10-fold cross validation (CV) scheme. In the outer loop, we divided the discovery dataset into 10 folds. After leaving one fold as the test set, the remaining nine folds were used as the training set. To minimise the bias arising from the imbalance between the number of patients with SSD and HCs, we conducted subsampling with undersampling simultaneously. We randomly sampled the same number (*N*=102) of HCs and SSDs from the training set, creating a subsample. During subsampling and undersampling, we matched the mean ages of HCs and SSDs. We fit the logistic regression model to the subsample while tuning a hyperparameter with an inner-loop 10-fold CV (for the detailed methodology, see **Supplementary Text 2**). By repeating random subsampling and fitting the model 10 times, we obtained 10 classifiers. We then predicted SSD probability for each participant in the test set. By applying the 10 classifiers built from a training set to a test set in each CV, we obtained the probability values for the participant as the classifier outputs. When the mean probability value was >0.5, we considered the participant’s class as SSD; otherwise, as HC. Finally, by repeating the above procedure 10 times in the outer loop, 100 classifiers were obtained.

We evaluated the performance of the classifiers using the following indices: an area under the receiver operating characteristic curve (AUC), accuracy, sensitivity, specificity, and Matthews’ correlation coefficient (MCC) (see **Supplementary Text 3** for details).

### Independent validation of LASSO classifiers

To assess the generalisability of the obtained classifiers, we tested them on the validation dataset. We applied the 100 classifiers to each participant in the validation dataset to compute diagnostic probability values for each participant (**Supplementary Figure 1b**). We classified a participant as having SSD if the average probability value was >0.5.

We also performed a statistical analysis of the classification performance for independent validation using a permutation test. We created 100 quasi-classifiers using the same procedure as for building genuine classifiers, with the participants’ classes permutated in the discovery dataset. We predicted the diagnosis in the same manner as mentioned above using the quasi-classifiers on the validation dataset for each permutation. By repeating random permutations 500 times and obtaining null distributions of the AUC and MCC, we evaluated the statistical significance of the true classifiers.

### Identifying important FCs for predicting diagnosis

We investigated which FCs were utilised to predict the diagnosis in each classifier by identifying the nonzero coefficients in LASSO. We counted the number of classifiers (out of 100) that selected each FC as an explanatory variable. We performed a permutation test to identify the most informative FCs from 71 631 FCs. Each time we randomly permutated the class labels of participants in the discovery dataset, we built 100 quasi-classifiers using 10-fold CV with 10-time subsampling, following the methodology described in the previous section (see ‘**Building LASSO classifiers’** section). We determined the maximum counts for which each FC was selected as a predictive explanatory variable using 100 quasi-classifiers for each permutation. This procedure was repeated 100 times, resulting in a null distribution of 100 values for the maximum selection count. An FC was considered significantly informative (‘important FC’) when the selection count in the genuine 100 classifiers exceeded the threshold that corresponded to *P* <0.05 in the null distribution.

### Voting classifiers

In addition to logistic regression with LASSO, we attempted to enhance the performance by introducing a voting method^33^. We incorporated support vector machine^34^, random forest^35^, light gradient boosting machine^36^, and multi-layer perceptron^37^ as representative algorithms. For each algorithm, we conducted CV, subsampling with undersampling, and training on the discovery dataset following the same procedure used to build the LASSO classifiers (for detailed methodology, see **Supplementary Text 4**).

To statistically compare the classification capability of the voting classifiers with that of the LASSO classifiers, we used the R programmes Compbdt and pROC. Using Compbdt^38^, we compared the two classifiers based on their sensitivity and specificity using the McNemar test. We used pROC to conduct DeLong’s test on the AUC. The significance level for all tests was set at *P* <0.05.

### Application of the classifiers to other psychiatric disorders

We assessed the specificity of the classifiers for SSD by applying them to data from participants with other mental disorders, MDD, ASD, and BP. Regarding the LASSO classifiers, we applied 100 classifiers to all the patients with these disorders from the discovery and validation datasets. If the output probability was >0.5, the participants were assumed to have SSD-like characteristics. Regarding the voting classifiers, we applied 500 classifiers to the same patients, and the participants were assumed to be SSD-like if over half (>250) of the classifiers predicted as such. The discovery dataset included patients with MDD, ASD, and BP, whereas the validation dataset only included patients with MDD. The outputs of the classifiers for HC and SSD were used for the comparison with MDD, ASD, and BP. Specifically, the output for test data in the 10-fold CV was used in the discovery dataset, and the output of 100 classifiers was used in the validation dataset.

To evaluate if the classifying results of patients with any of the other disorders (MDD, ASD, BP) had similarity to HC, SSD, or neither, we conducted a two-sided binomial test (significance level, *P*<0.05). We also assessed whether each probability density curve was differently distributed using the two-sample Kolmogorov–Smirnov test. We conducted this test for every combination of HC, SSD, MDD, ASD, and BP; thus, 10 combinations were obtained in total. The level of significance was *P*<0.05/10=0.005 (Bonferroni-corrected).

### Prediction of clinical scores using important FCs

Another objective of this study was to determine the extendibility of the trait marker to the state or disease stage. In the first part of this investigation, we attempted to predict the scores of two clinical scales with important FCs *in the aggregate* so we could aid clinical assessment and estimate the extent to which our biomarker could function as a trait and/or state marker using the PDI and PANSS total scores. These scales have been widely used to assess delusional thinking and psychotic syndrome^39^. PDI may reflect trait^20^ and state^40^, whereas the PANSS may reflect the overall symptom severity of psychosis at that time point (state)^41^.

We predicted the scores on the clinical scales (PDI and PANSS total scores) using important FCs as explanatory variables. We applied a nested 10-fold CV scheme to the discovery dataset (only SSD participants with available target scale scores) to build prediction models (**Supplementary Figure 2a**). In the outer loop, we divided the discovery dataset into 10 folds. After leaving one fold as the test set, we used the remaining nine folds as the training set. We fitted the linear regression model to the training set (the *LinearRegression* module in scikit-learn version 0.24.1). In the inner loop, we searched for the most suitable number of important FCs to be used as features to avoid overfitting (for the detailed methodology, see **Supplementary Text 5 and Supplementary Figure 2b**). We predicted the score for each participant in the test set using a linear regression model with the best number of features for the fold. Using 10-fold CV, we obtained 10 models with different numbers of features. For the validation dataset, we averaged the outputs from the 10 models to create the final predicted score (**Supplementary Figure 2c**). At any step of this analysis, the predicted value was adjusted within the range of the scale score (if the prediction was lower than the lowest possible value, it was adjusted to the lowest possible value, and vice versa).

We conducted a permutation test to assess the evaluation metrics statistically. Specifically, we permutated clinical labels (diagnosis and symptom scores) against FC values, created quasi-models following the same procedure as the genuine models, and repeated these steps 500 times to obtain a null distribution of the evaluation metrics. Differences were considered statistically significant if each evaluation metric for the genuine models was better than the cut-off value of *P*=0.05 of the null distribution.

### Separately identifying FCs associated with the state

Next, we explored the roles of individual FCs. To identify the FCs associated with state (i.e. symptom scale scores), we performed multiple regression analyses on each of the important FCs. We selected PANSS as the state index because it dynamically changes with treatment and has been used as a representative scale for the state of schizophrenia. All HC participants and patients with SSD having data on PANSS scores across the discovery and validation datasets were included in the analysis. We examined two models of the PANSS: the original three-factor model^19^ composed of positive, negative, and general pathological factors and the five-factor model^41^ composed of positive, negative, disorganised, excited, and depressed factors. Because the general pathological factor was not categorised as a specific symptom dimension, we used only positive and negative factors as explanatory variables for the three-factor model.

Owing to the exploratory nature of this analysis, we converted the PANSS scores into explanatory variables in two ways: by min–max normalisation and binary dummy variables. The formula to fit was the same for both methods:

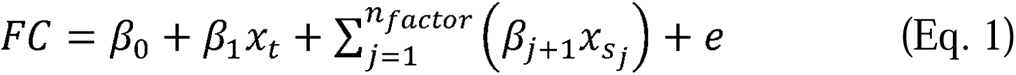

where *x_t_* represents the trait or diagnosis (HC: 0, SSD: 1), 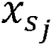 represents the existence of symptoms for the *j*th factor of the PANSS, and *e* represents a random error.

#### Conversion 1. Min–max normalisation of the scores

The value 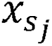 was determined using the following formula:

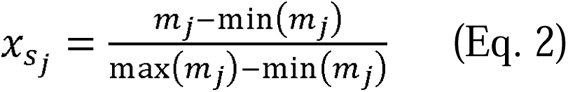

where *m_j_* represents the participant’s average score in the *j*th factor of the PANSS and *min*(*m_j_*) (or *max*(*m_j_*)) represents the minimum (or maximum) average score of the *j*th PANSS factor across all patients with SSD with available PANSS subscore data.

#### Conversion 2. Binarising the scores

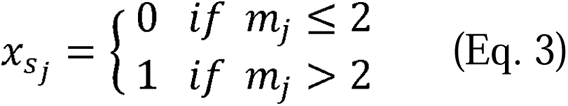

The value 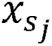 was determined as in Eq.3 as three points or more is considered pathological in PANSS rating.

All *s_j_* for HC participants were assumed to be zero. In the regression analysis for each FC, the FC was regarded as a potential state marker when any of the coefficient estimates for PANSS factor (*β*_(*j*+l)_, *j*; = 1,2,… *n_factor_*) was nonzero at a significance level *P* <0.05 with a one-sample *t*-test. We reported FCs as being significant only when the state’s coefficient concerned (*β*_(*j*+l)_) was of the same sign as *w* of the FC in terms of consistency with the underlying LASSO classifiers.

### Identifying FCs associated with the disease stage

Finally, we constructed another multiple regression model to identify FCs associated with the disease stage of SSD, which could be referred to as ‘staging’ markers. Following a pre-existing definition^42,43^, we divided the SSD group into an early-stage psychosis subgroup (DOI of <5 years) and a chronic-stage subgroup (DOI of ≥5 years). We formulated a regression model similar to that described in the previous section:

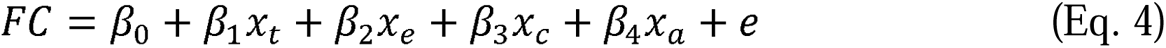

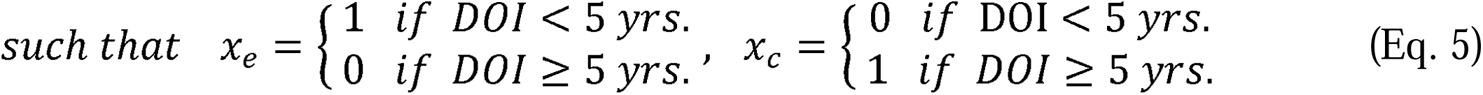

where *x_e_* represents the early stage of SSD, *x_c_* represents the chronic stage, and *x_t_* and *e* were defined in the same way as in Eq. 1. The value *x_a_* the age of the participant, was introduced as a covariate because the DOI was supposed to correlate with age. All HC participants and patients with SSD with DOI data across the discovery and validation datasets were included in the analysis. Using this model, we categorised important FCs into six groups based on which trait, early/chronic stage, was significantly associated: (1) trait only, (2) early stage only, (3) chronic stage only, (4) trait and early stage, (5) trait and chronic stage, and (6) all three. In terms of consistency with our classifiers and within the coefficients, every coefficient estimate concerned must be of the same sign as *w*.; statistical significance was determined at the level *P*<0.05, with a one-sample *t*-test for each coefficient estimate. Considering the relative instability of the model fit due to singularity, we adopted the bootstrap method (1,000 iterations) to count the number of times a certain FC was sorted into each category.

## RESULTS

### Datasets

Fifty-one participants whose scrubbed rs-fMRI volumes exceeded +3 standard deviations (SDs) were excluded from all datasets. Therefore, 1,015 participants in the discovery dataset and 683 participants in the validation dataset were included for further analyses. Through the scrubbing process, 9.2±17.9% (mean±SD) of whole volumes per rs-fMRI scan were removed across all datasets.

### Performance of the LASSO classifiers

Within the discovery dataset, classification accuracy was 79.6%, with an AUC of 0.876. Sensitivity, specificity, and MCC were 81.5%, 79.2%, and 0.484, respectively. The density curve of HCs and patients with SSD is shown in **Figure 2a**, where a patient with a predicted probability of >0.5 was classified as a patient and vice versa. The curves for each site are shown in **Figure 2b**.

**Figure 2.**
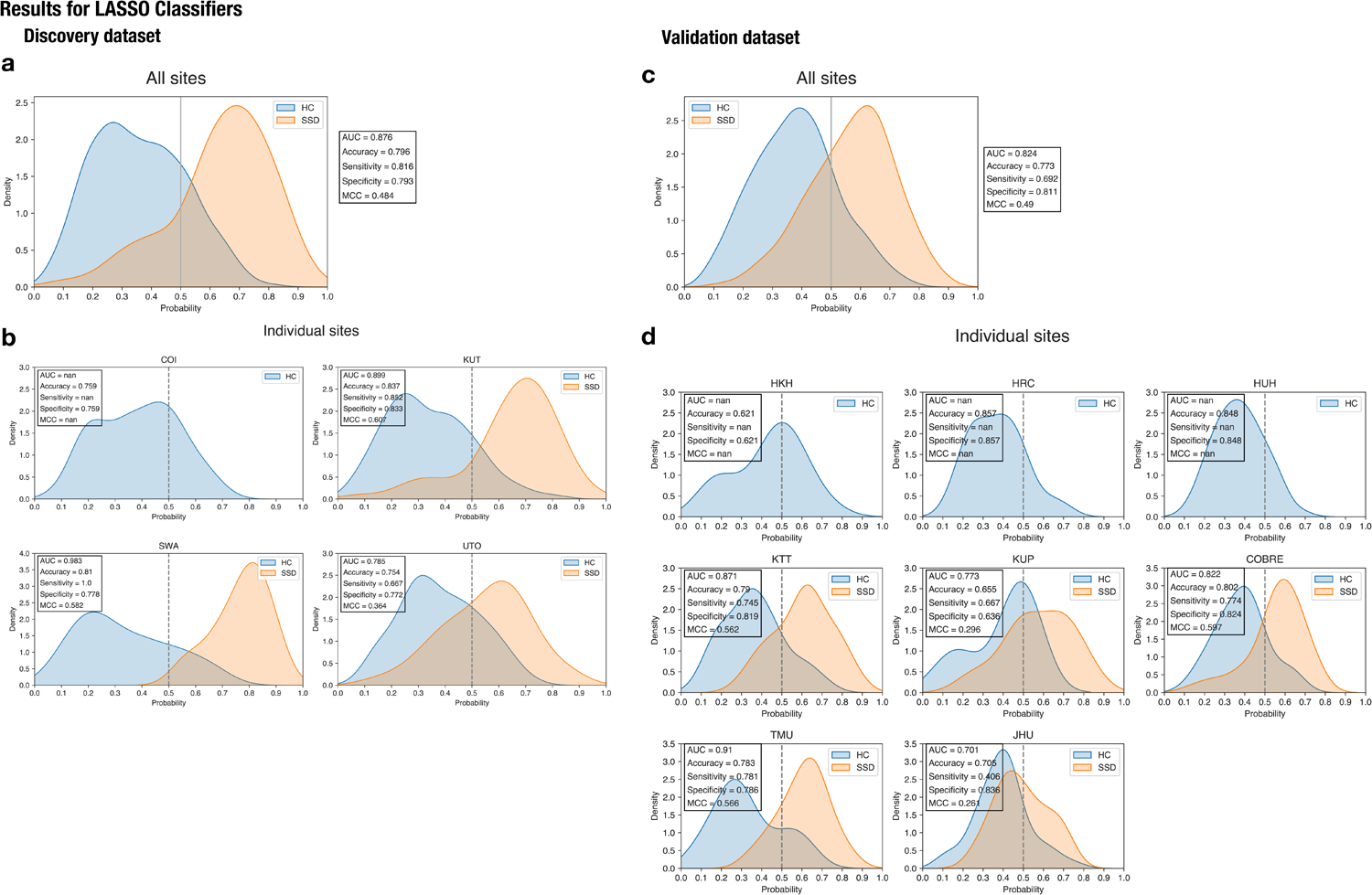
Probability density curves based on LASSO classifiers. **(a)** Results for all the sites combined in the discovery dataset. **(b)** Results for individual sites in the discovery dataset. **(c)** Results for all the sites combined in the validation dataset. **(d)** Results for individual sites in the validation dataset. As four sites in Hiroshima (COI, HKH, HRC, and HUH) did not have any patients with SSD, these sites have a curve for HCs only. HC, healthy control; SSD, schizophrenia spectrum disorder; AUC, area under the curve; MCC, Matthews’ correlation coefficient; LASSO, least absolute shrinkage and selection operator; COBRE, Centre of Biomedical Research Excellence.

Within the validation dataset, the classifiers distinguished patients with SSD from HCs with 77.3% accuracy and an AUC of 0.824, similar to the results from the discovery dataset (**Figure 2c, d**). Sensitivity, specificity, and MCC were 69.2%, 81.1%, and 0.490, respectively. A permutation test revealed that the AUC and MCC were significantly high (*P*<0.001). At JHU, the classifiers correctly distinguished HCs in most cases (specificity, 83.6%), but not patients with SSD (sensitivity, 40.6%). The probability density curve of SSD was closer to that of HC (*P*=0.37, two-sided binomial test), although the distribution of the two curves was significantly different (*P*=0.013, two-sample Kolmogorov–Smirnov test), suggesting that patients with early-stage SSD at JHU fell between chronic-stage SSD and HC.

### Important FCs for predicting diagnosis

At *P*<0.05 in the permutation test, the FCs selected by ≥17 classifiers were deemed important. Ultimately, we identified 47 important FCs, as shown in **Figure 3a, b** and detailed in **Supplementary Table 4**.

**Figure 3.**
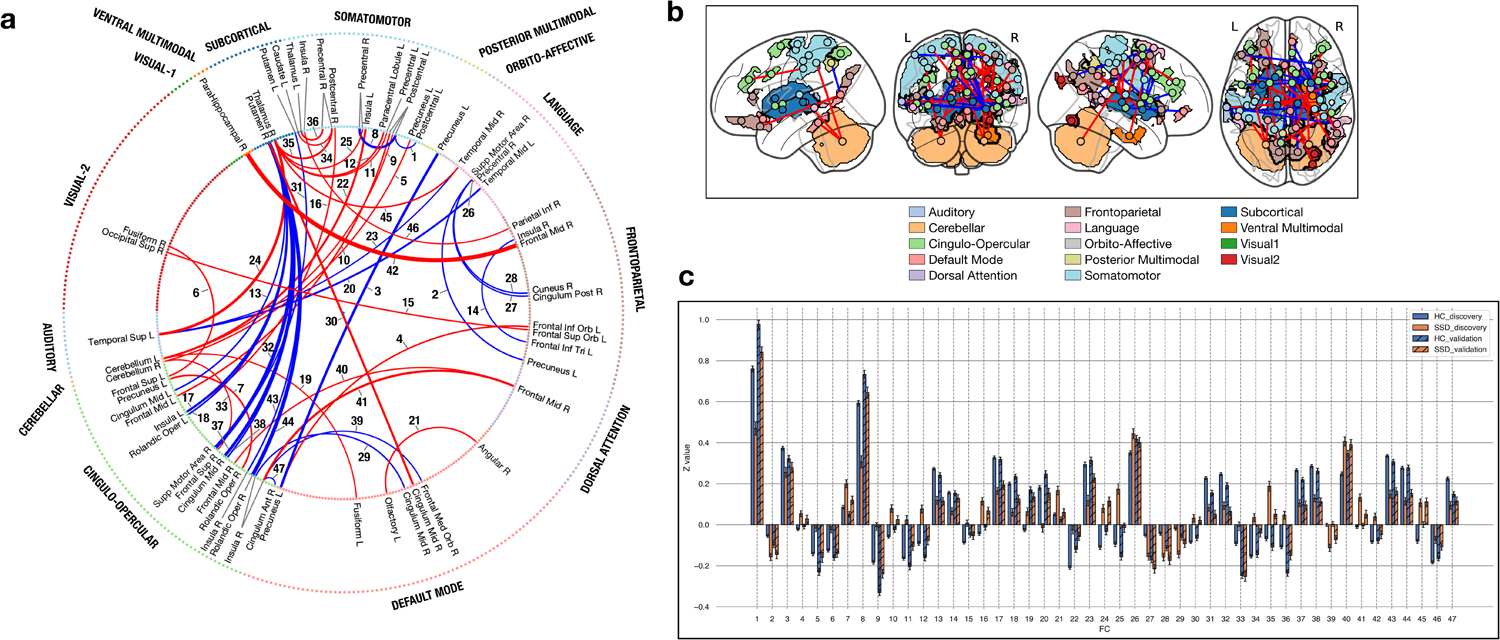
Important FCs (P<0.05) in diagnosis prediction by LASSO classifiers. **(a)** Each node on the inner circle corresponds to an ROI. The line width of the FC shows how many times it was selected by the classifiers, and the line colour denotes the direction to which it contributes to the logistic regression model (red means that the higher the FC value is, the more likely the classifier’s output is to be SSD; blue means a lower FC value for a higher likelihood of SSD). **(b)** The 47 important FCs were projected onto glass brains. The colours of the ROIs in relation to each intrinsic brain network and the red/blue line colours correspond to those in (a). **(c)** The mean FC values (Z-score, on the ordinate) of the 47 important FCs for the HC and SSD groups are shown as a bar plot for the discovery and validation datasets. Error bars represent standard error. The FC numbers on the abscissa correspond to those in (a) and **Supplementary Table 4**. FC, functional connectivity; HC, healthy control; SSD, schizophrenia spectrum disorder; ROI, region of interest; LASSO, least absolute shrinkage and selection operator.

Furthermore, we investigated the detailed patterns of FC value differences between the HC and SSD groups and their reproducibility across the two datasets by plotting the mean values for 47 FCs, facilitating a comparison between the datasets (**Figure 3c**). The relationship between the mean FC values of the HC and SSD groups was maintained in the validation dataset for 44 of 47 FCs.

### Voting classifiers

**Supplementary Figure 3a, b** displays the probability density curve of the voting classifier’s output through 10-fold CV in the discovery dataset.

Independent validation for all sites in the validation dataset is presented in **Supplementary Figure 3c, d**. We conducted statistical tests to compare the performance of the voting classifiers with that of the LASSO classifiers. Sensitivity was significantly higher for the voting classifiers than for the LASSO classifiers (*P*<0.001, McNemar test, two-sided). However, specificity showed no significant difference between groups (*P*=0.192, McNemar test, two-sided). The AUC of the voting classifiers was 0.841, which was not significantly different from that of LASSO classifiers (*P*=0.11, DeLong’s test).

### Supplementary analyses on classifiers’ characteristics

LASSO and voting classifiers were built on subsamples in which the mean ages of the HC and SSD groups were matched to mitigate the confounding effect of age. Furthermore, we evaluated whether the classifiers’ performance was influenced by confounding factors (see **Supplementary Text 6** and **Supplementary Table 5**). Consequently, the performance of voting classifiers could be partly influenced by age but not by other potential confounding factors. In contrast, the LASSO classifiers were unaffected by any potential confounding variables.

We also investigated how the classification performance varied based on disease severity in the validation dataset. Following a previous report^44^, we identified subgroups with different severities based on PANSS total scores: mild (PANSS≤58, *N*=55), moderate (58<PANSS≤75, *N*=36), marked (75<PANSS≤95, *N*=20), severe (95<PANSS≤116, *N*=1), and most severe (116<PANSS, *N*=0). Except for the severe and most severe subgroups, each of which contained only one or no participants, the sensitivity by subgroup was highest in the marked subgroup, followed by the mild and moderate subgroups (**Supplementary Table 6**). This order of performance was consistent for the LASSO and voting classifiers, but the disparity among the subgroups decreased for the voting classifiers compared with the LASSO, suggesting that voting classifiers can predict classes with less imbalance across different disease severity levels.

### Classifier specificity for SSD

The LASSO classifiers revealed that the patients with ASD and BP did not exhibit high- or low-SSD-like characteristics. However, patients with MDD were less SSD-like (*P*=2.24×10^−7^), resembling the HCs (**Supplementary Figure 4, Supplementary Table 7**). Voting classifiers showed a similar pattern. These results suggest that both classifiers exhibited high specificity for SSD.

Moreover, we investigated whether each probability density curve was distributed differently. The LASSO and voting classifiers revealed that the curve of each non-SSD disorder had a significantly different distribution from those of HC and SSD in the discovery and validation datasets (**Supplementary Table 8**). Furthermore, the curves for any two non-SSD disorders showed no significant differences.

### Prediction of clinical scale scores using important FCs

Regarding PDI total scores, the results of 10-fold CV in the discovery dataset were acceptable (*r*=0.231, 95% confidence interval [CI]=[0.00101–0.438], *P*=0.0492, two-sided, mean absolute error [MAE]=40.0). Weak correlations between the actual and predicted scores were observed in the validation dataset (*r*=0.331, 95% CI=[0.0609–0.556], *R*^2^=0.110, *P*=0.0177, two-sided, MAE=55.5) (**Figure 4a**). A permutation test was conducted to objectively assess whether the evaluation metrics were satisfactory. The correlation coefficients were significantly high (*P*_perm_=0.01) and MAE significantly low (*P*_perm_=0.018).

**Figure 4.**
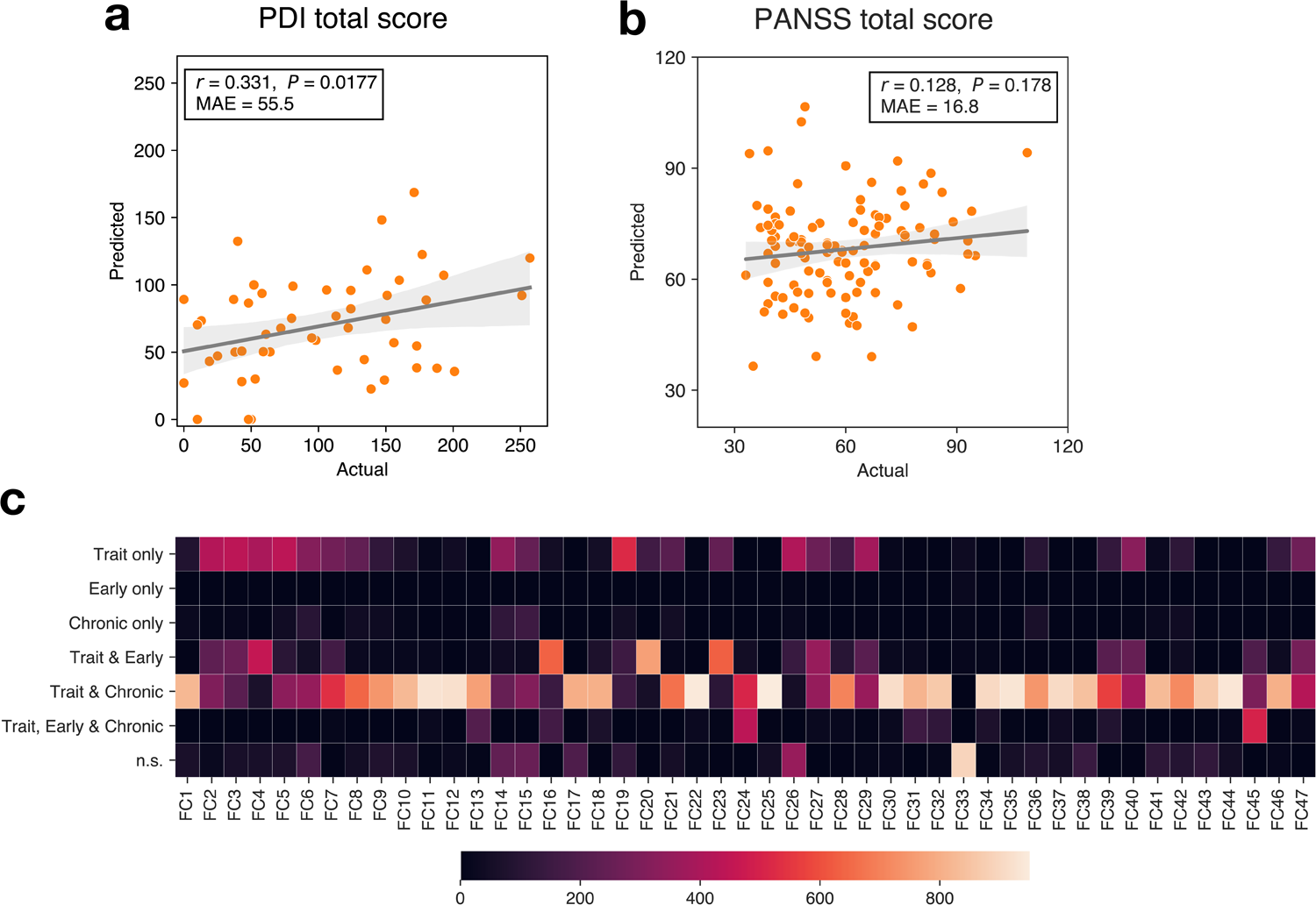
Trait, state, and staging marker analyses. **(a, b)** Prediction results of PDI total and PANSS total scores. The relationship between the predicted scores (on the ordinate) and actual scores (on the abscissa) for the validation dataset is shown in a scatter plot. The grey translucent band represents the 95% confidence interval of the regression line. **(a)** PDI total scores. **(b)** PANSS total score. **(c)** Individual FCs associated with disease stage. This heat map shows the results of the bootstrap method, where the number in the colour bar indicates the number of times the FC was sorted into a category out of 1,000 iterations. n.s., not significant (*P*≥0.05 for all three explanatory variables or any of the coefficients’ signs were inconsistent with the average weight in LASSO classifiers); MAE, mean absolute error; PDI, Peters et al. Delusion Inventory; PANSS, Positive and Negative Syndrome Scale; FC, functional connectivity; LASSO, least absolute shrinkage and selection operator.

Concerning PANSS total scores, the 10-fold CV in the discovery dataset resulted in no correlation between actual and predicted scores (*r*=−0.0065, 95% CI=[−0.199–0.186], *P*=0.948, two-sided, MAE=17.5). The model did not predict scores in the validation dataset either (*r*=0.128, 95% CI=[−0.0588–0.306], *R*^2^=0.016, *P*=0.178, two-sided, MAE=16.8) (**Figure 4b**).

In summary, we predicted the PDI total score more efficiently than the PANSS total score, suggesting that, collectively, the important FCs may function as trait and state markers.

### Individual FCs associated with PANSS factors

In total, 17 FCs were significantly associated with the PANSS factors. Using the three-factor model of PANSS, we identified significant associations in FCs #45 (R.TPOJ1 and R.Thalamus, with the PANSS positive factor) and #8 (L.1 and R.3b, with the PANSS negative factor), regardless of the score conversion method (**Table 2**, **Supplementary Table 9**). With the five-factor model of PANSS, we found that the significantly associated FCs, regardless of conversion method, were FCs #18 (L.FOP1 and R.Putamen), #41 (R.6a and R.PoI1), #43 (R.FOP4 and R.Putamen) with the excited factor, and FC #27 (R.RSC and R.SFL) with the depressed factor.

**Table 2.**
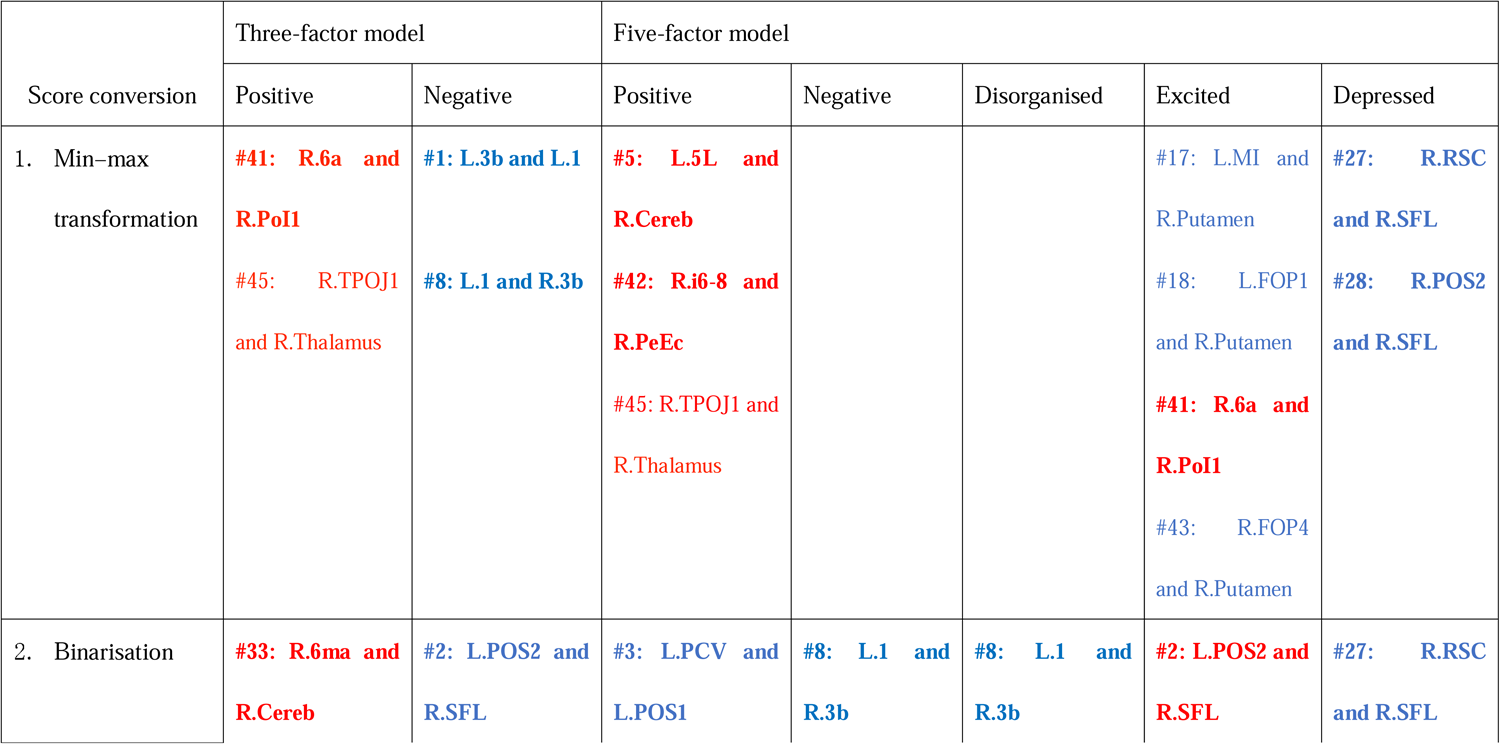
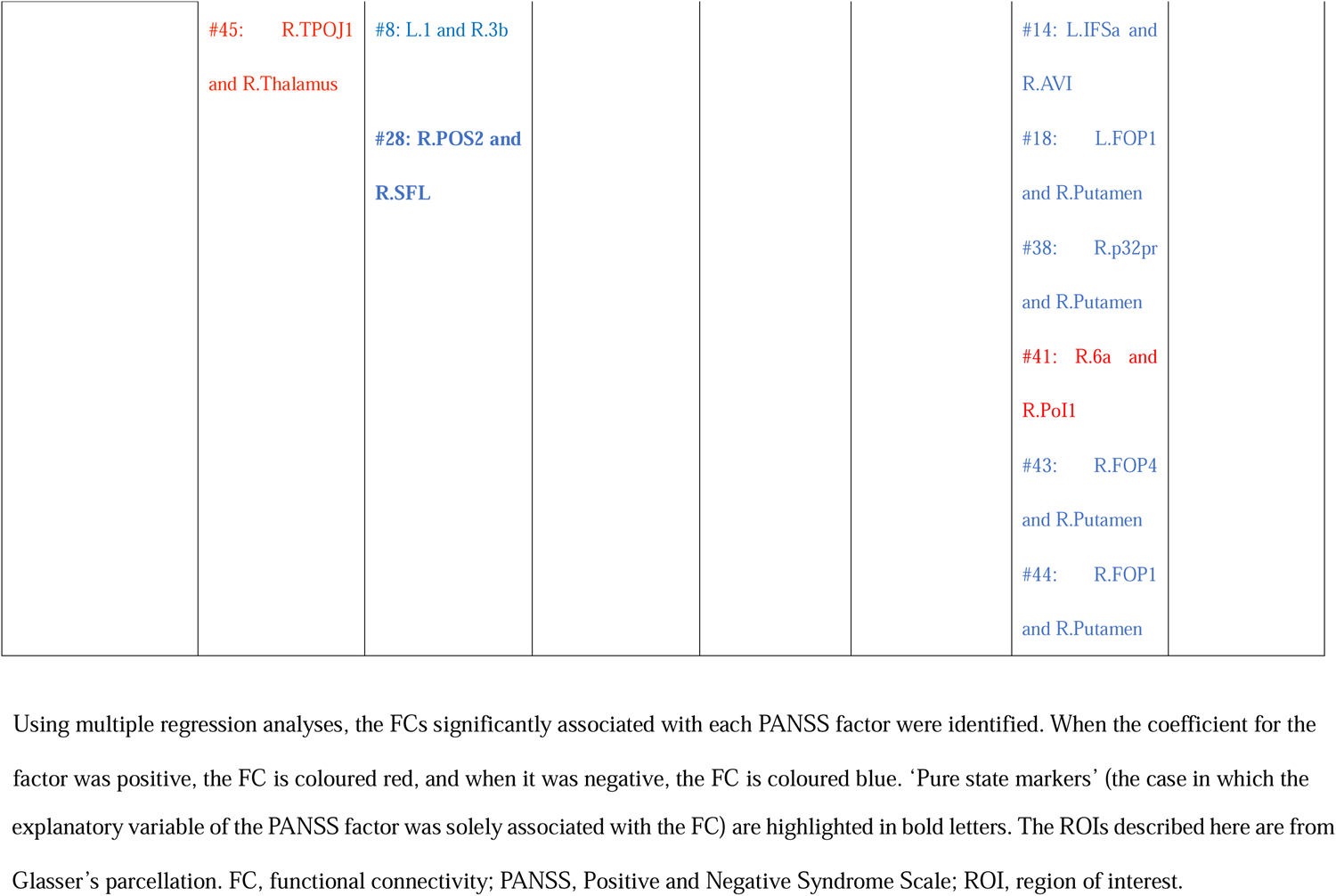
Individual FCs significantly associated with PANSS factorial scores.

When the coefficient estimate was significant only for a state (not for the trait), such FCs were assumed to be ‘pure’ state markers. We found nine FCs (**Table 2**). Among these FCs, FC #27 (R.RSC and R.SFL) was the only one consistently identified as a ‘pure’ state marker, regardless of conversion method.

### Individual FCs associated with disease stage

Figure 4c presents the results of categorising important FCs into subgroups associated with trait (diagnosis), the early stage, or the chronic stage of SSD using the bootstrapping method (1,000 iterations). We assumed that an FC belonged to a certain category with consistent regression results for over half (>500) of the iterations. Subsequently, the important FCs were categorised most frequently as ‘trait and chronic stage’ (61.7%), followed by ‘trait and early stage’ (6.4%), ‘trait only’ (2.1%), and ‘trait, early, and chronic stage’ (2.1%). No FCs were categorised as ‘early stage only’ or ‘chronic stage only,’ indicating that we did not identify any pure staging markers.

## DISCUSSION

We developed rs-FC classifiers for SSD based on harmonised multicentre data and validated it using large-sample independent data. This study also aimed to maximise the potential of the rs-FC biomarker to function as a state marker and a staging marker.

The accuracy of the identified diagnostic marker was approximately 80% for the discovery dataset. It demonstrated comparable performance in an external validation with seven international cohorts; this presents a sharp contrast to previous studies^10^. This achievement was possible through bi-directional (prospective and retrospective) harmonisation and with optimised machine learning method. Against the concerns on session variability of rsfMRI, we reduced the variability and improved generalisability to independent validation cohorts through spatial averaging of 100 classifiers, each analysing tens of FCs. Furthermore, a previous study^45^ revealed that test–retest reliability was acceptable with the same methodology as demonstrated in this study.

The LASSO and voting classifiers exhibited distinct performance characteristics. Voting classifiers demonstrated superiority in sensitivity and a more balanced profile over LASSO classifiers, with the accuracy, sensitivity, and specificity falling within a narrow range (LASSO: 69.2–81.1%, voting: 74.7–78.8%) in independent validation.

We identified important FCs that significantly contributed more frequently to SSD classification in LASSO classifiers. Based on AAL, the putamen, insula, thalamus, and cingulum were among the top ROIs most frequently found in the important FCs (**Supplementary Table 10**). In schizophrenia research, these are consistently implicated in grey matter volume reduction^46^ or FC abnormality^47,48^. Moreover, these ROIs were associated with the cortico-striatal-thalamic-cortical loop and salience network, both recognised for their pivotal roles in the psychopathology of SSD^49^. In the context of FC, hypoconnectivity between the putamen and anterior cingulate cortex (FC #13, #30, #37, #38) in the SSD group aligns with a previous report^50^, where greater ACC–putamen connectivity predicted better treatment response. Conversely, in the SSD group, the thalamus exhibited increased connectivity with various cortical ROIs (FC #11, #12, #24, #25, #35, #36, and #45), corresponding with a report^51^ on thalamocortical connectivity in schizophrenia, implying disrupted information filtering in SSD, consistent with the literature^52^. Therefore, these important FCs aptly reflected the neural correlates of schizophrenia and were considered trait markers of SSD.

We observed that our classifiers exhibited high specificity for SSD, whereas other diagnoses (MDD, ASD, and BP) did not show specificity for HC or SSD. These psychiatric disorders share several characteristics with SSD in the alteration pattern of brain function^53–55^. Moreover, these disorders have similar phenotypes (e.g. cortical thickness) and genotypes^56^. Thus, our biomarkers may, to some extent, reflect neural changes common to psychiatric disorders^57^.

The second objective of this study was to dissect the biomarker into a ‘trait marker’ and other components. Using aggregated important FCs, we achieved an acceptable level of prediction for the PDI but not for the PANSS, indicating that aggregated FCs were more strongly associated with traits. Few groups have reported successful prediction of symptoms using FC^58,59^; however, these studies lacked external validation. In this study, the predicted PDI total score was significantly correlated to the actual score in the discovery and validation datasets.

Using multiple regression analysis, we identified state markers individually. Using the three-factor model of the PANSS, we observed a significant association between positive scores and FC #45 and negative scores and FC #8, irrespective of the score conversion method. Evidence suggests that FC #45 significantly correlates with positive symptoms^60^. FC #8 also showed a significant association with the negative and disorganised factors of the five-factor model, which could be linked to the finding that interhemispheric connectivity of the precentral and postcentral gyri were negatively correlated with the PANSS total score^61^.

We discovered interesting overlaps of ROIs in the potential state marker FCs. Concerning negative and depressed factors, sensorimotor areas were noticeable. Specific functional alterations in these ROIs related to neurological soft signs (NSSs) have been reported^62^. Moreover, NSSs correlate with PANSS negative scores and depression scale scores^63^. The excited factor seems to involve the putamen, insula, Rolandic operculum, and middle cingulum, regions likely to be associated with aggression^64^. The Rolandic operculum and middle cingulum are related to aggression via disruption of the cognitive control network^65^.

Neuromodulation can be a novel psychiatric treatment, and one of the promising techniques is neurofeedback (Nef). There is accumulating evidence on the therapeutic effect of Nef targeting FCs^66,67^. Our findings on state markers will benefit future FC-Nef in target selection.

Although we did not discover any pure staging markers, our results provide a compelling argument for ‘trait and early stage’ markers exemplified by FCs #16, #20, and #23. FCs #20 and #23 represented connections between regions around the superior temporal sulcus (STS) or gyrus (STG). A task-based fMRI study on working memory showed attenuated activity in the STG in patients with early-stage psychosis compared with HCs^68^. Moreover, the cortical thickness of the insula (an ROI in FC #16) and the STS region are reduced in early-stage psychosis^69^. These suggest that our biomarker partly include staging marker. Additionally, the fact that the probability curve for patients with early-stage psychosis at JHU lies in the middle between the HC and SSD groups may also support this argument.

This study has some limitations. First, the participants with SSD in our study were mostly medicated; hence, the applicability of our results to drug-naive patients cannot be guaranteed. Second, we only confirmed that our classifiers distinguished patients with SSD from healthy individuals, not from individuals with other psychiatric disorders exhibiting psychotic symptoms. Further studies are required to test the classifiers for these disorders to maximise the clinical applicability.

In conclusion, we developed robust and clinically usable classifiers for SSD using a combination of cutting-edge strategies. While constructing the classifiers, we identified FCs that may play key roles in SSD pathophysiology. We also demonstrated that these ‘important FCs’ had multiple functions as SSD trait, state, or staging markers. Our findings shed new light on the early diagnosis of SSD and the selection of targets for neuromodulation.

## Data Availability

All data produced in the present study are available upon reasonable request to the authors.

## Acknowledgements

We thank Editage (www.editage.com) for the English language editing.

## Author contributions

T.K., A.Y., M.K., and H.T. designed the study. T.K., Y.Y., Y.K., N.O, K.K., M-C.H., A.S., T.M., J.M., and H.T. recruited participants for the study and collected their clinical and imaging data. T.K. and A.Y. performed the data analysis. T.K., A.Y., and J.M. wrote the original draft of the manuscript. T.K., A.Y., Y.Y., Y.K., N.O, K.K., M-C.H., A.S., J.Y., O.Y., T.M., J.M., M.K., and H.T. reviewed and revised the manuscript.

