## Supplementary figures and images for "Generalisable functional imaging classifiers of schizophrenia have multifunctionality as trait, state, and staging biomarkers"

### Supplementary Figure 1

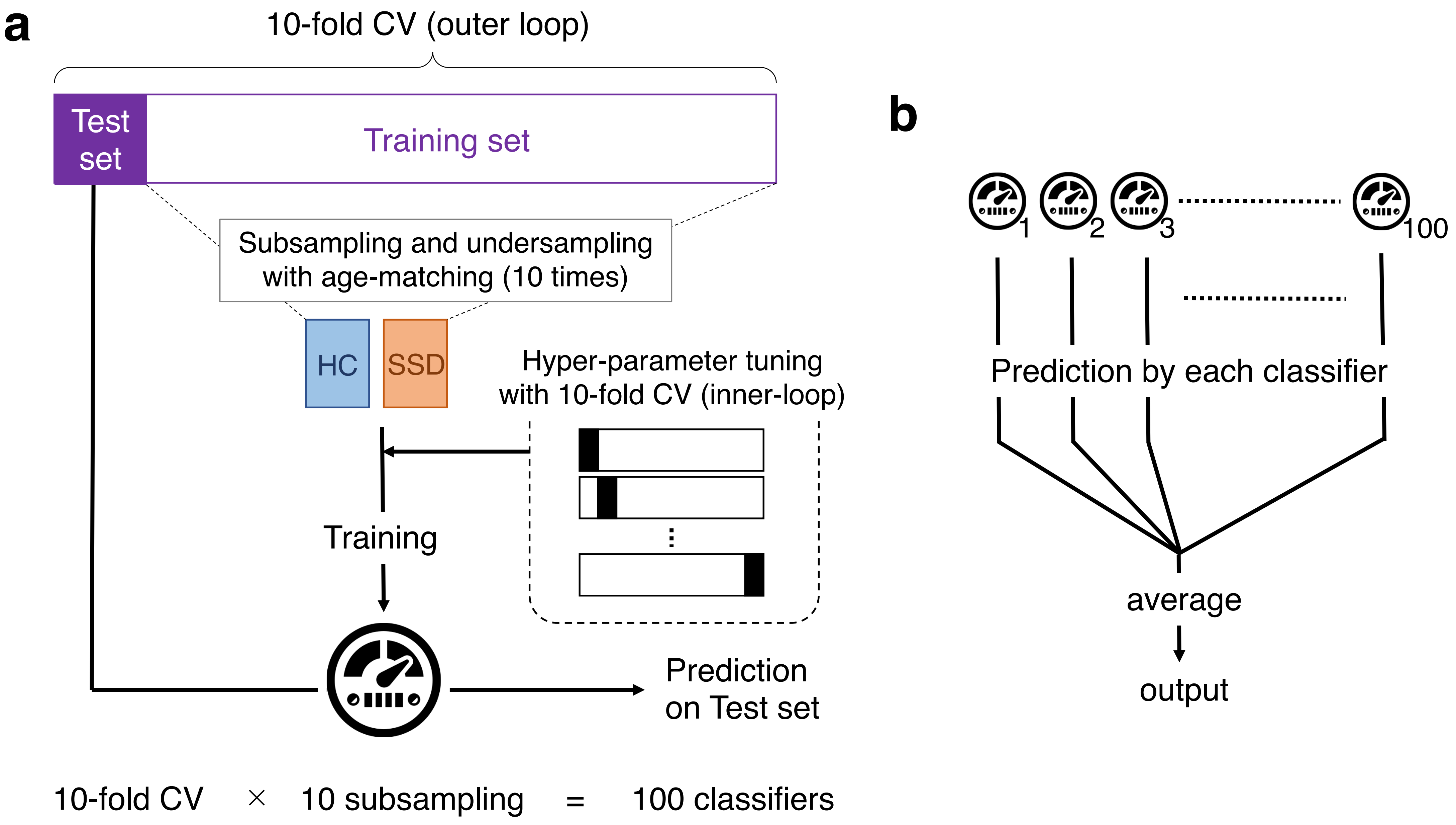

### Supplementary Figure 2

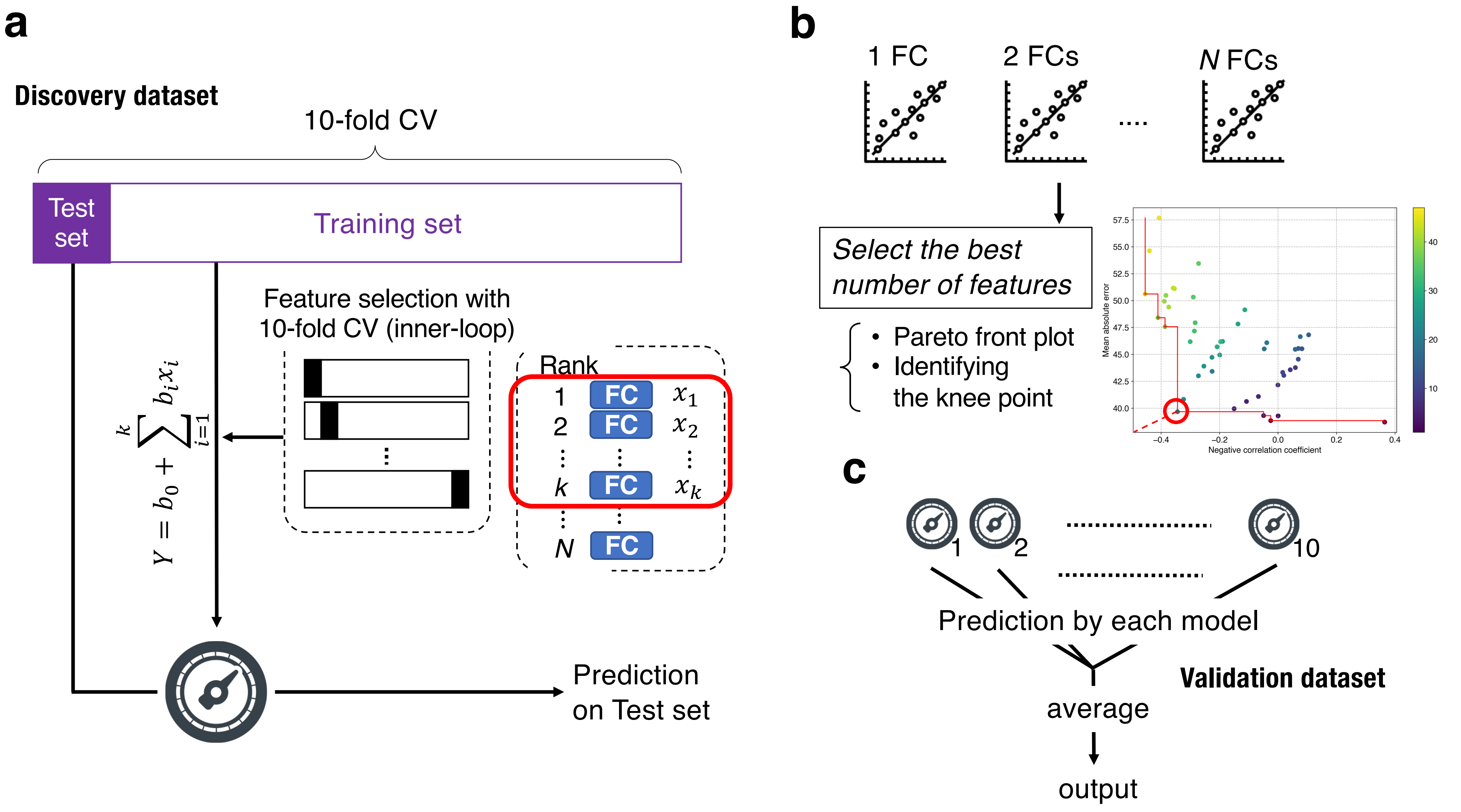

### Supplementary Figure 3

#### Results for Voting Classifiers

##### Discovery dataset

##### Validation dataset

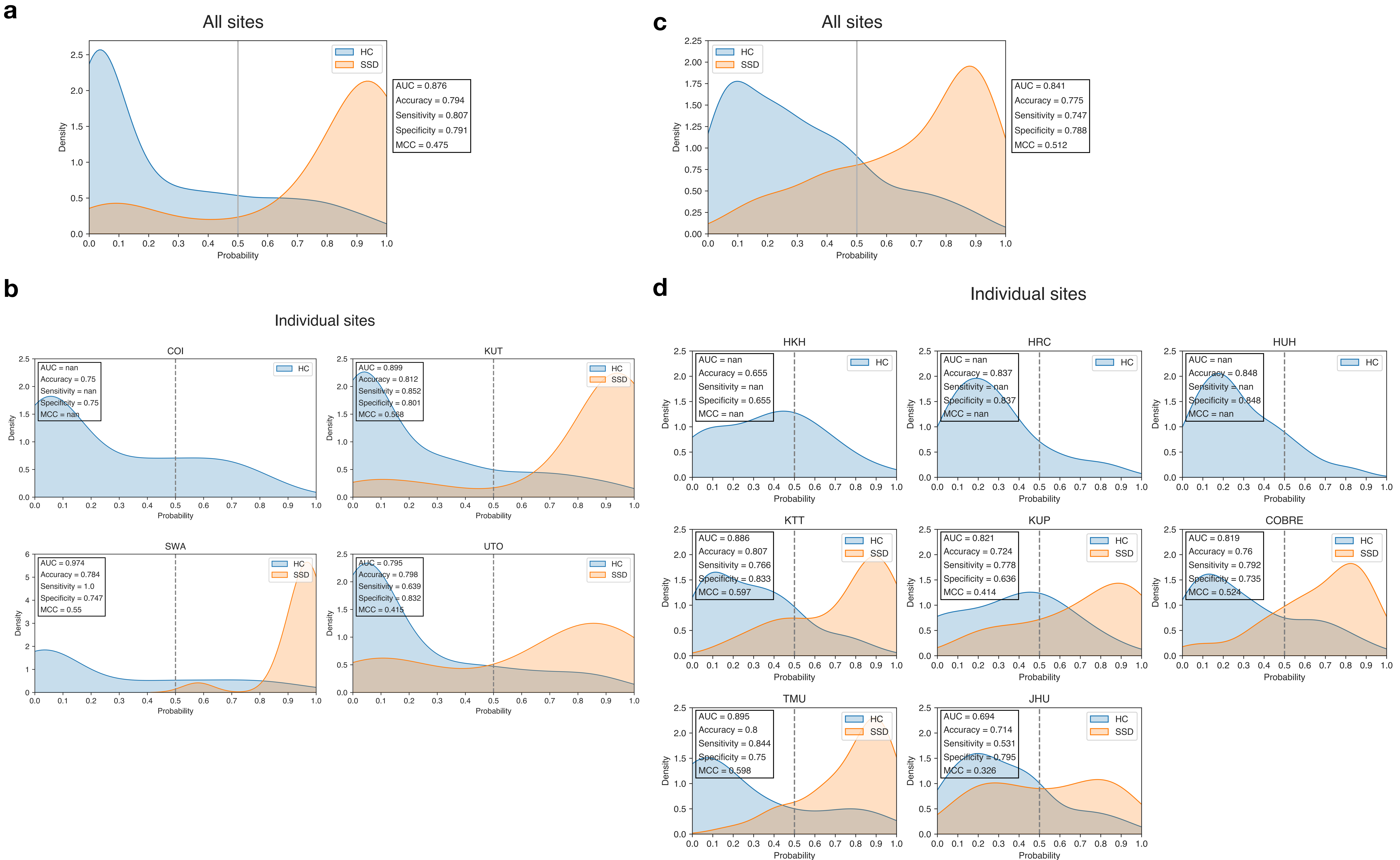

### Supplementary Figure 4

##### a LASSO Classifiers

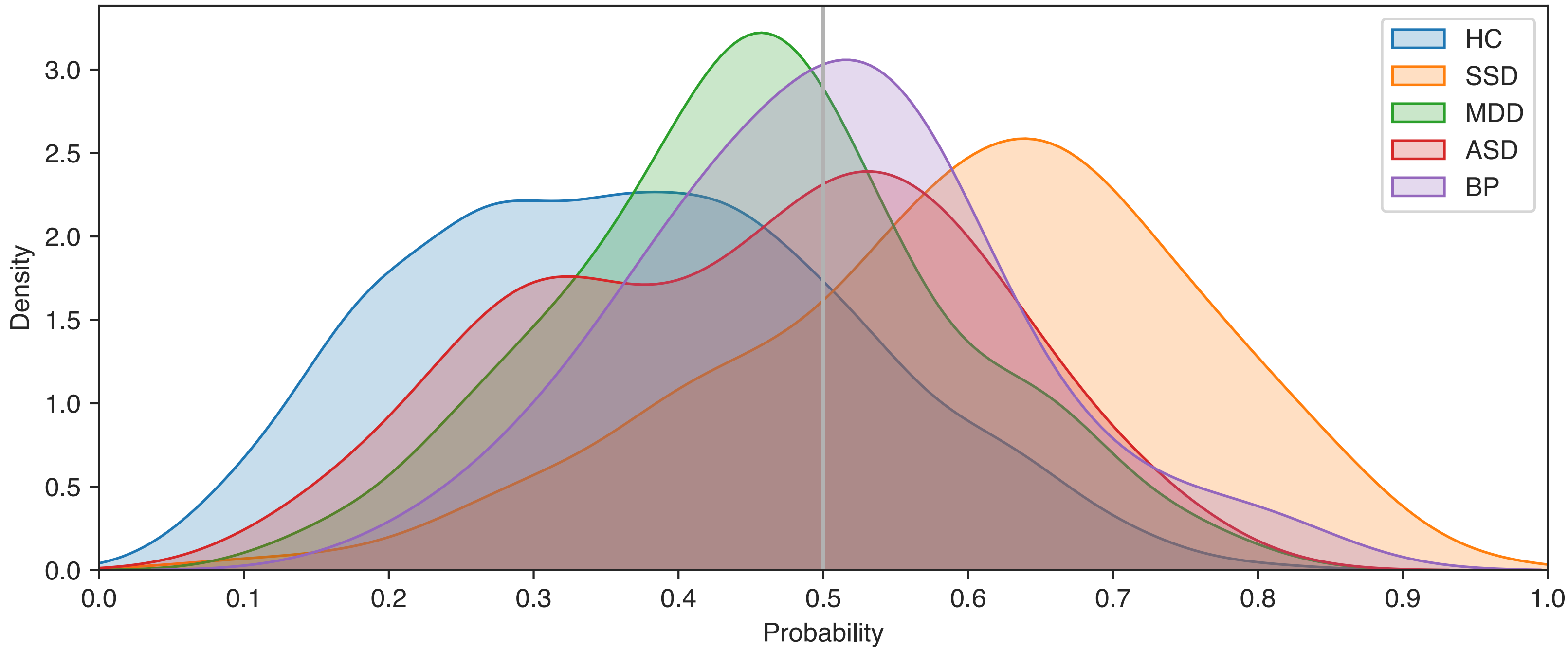


##### b Voting Classifiers

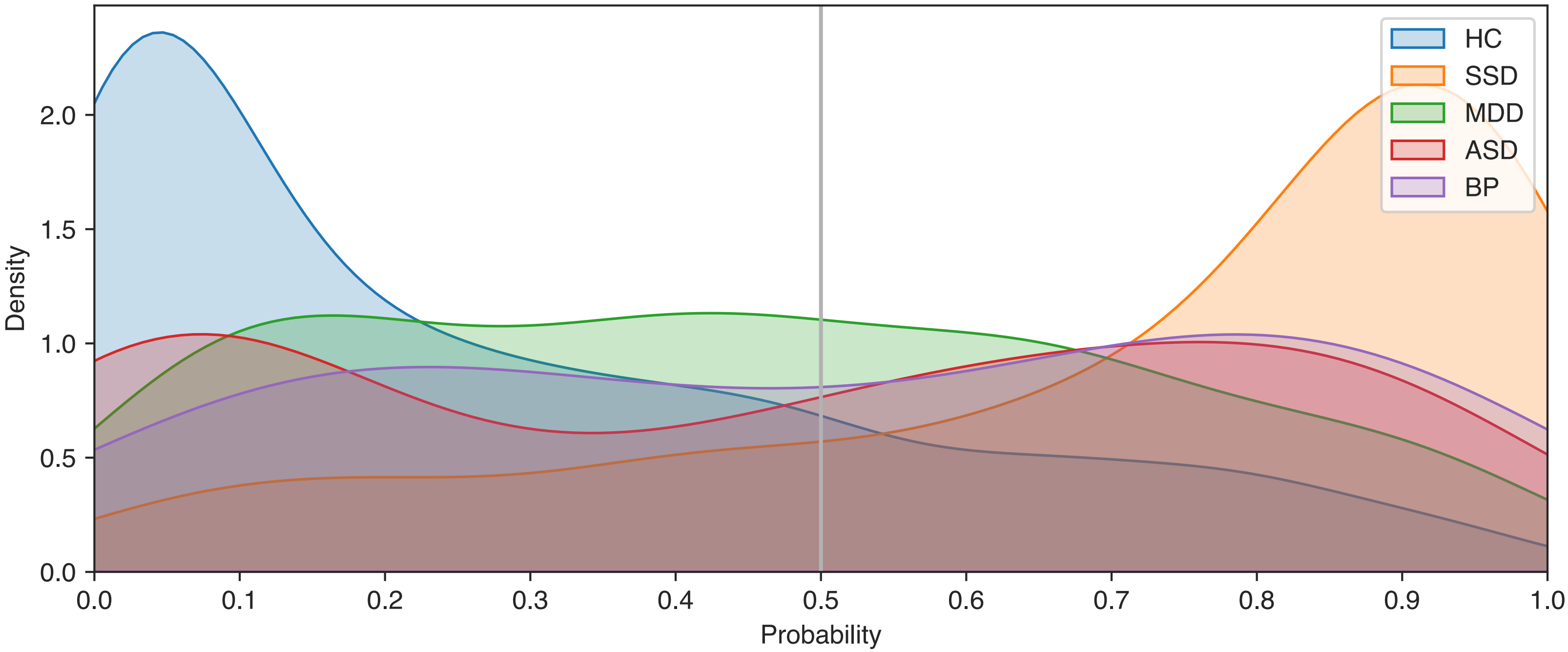
