## Supplementary Figure Legends for "Generalisable functional imaging classifiers of schizophrenia have multifunctionality as trait, state, and staging biomarkers"

**Supplementary Figures**

**FIGURE LEGENDS**

**Supplementary Figure 1.** **Scheme of the procedures for building classifiers**

**(a)** Workflow chart of the nested 10-fold CV applied to the discovery dataset. By repeatedly subsampling and undersampling from a training set ten times, we obtained ten subsets for each CV fold. Ultimately, 100 classifiers were obtained. **(b)** The resulting 100 classifiers were applied to the validation dataset. The average value of the classifier output was the final output. CV: cross-validation, HC: healthy control, SSD: schizophrenia spectrum disorder.

**Supplementary Figure 2. Scheme of the procedures for predicting symptom scores**

**(a**) Workflow chart of fitting models in the nested 10-fold CV on the discovery dataset. The top-ranked FCs of a particular number (*k* ∈ 1, 2, ..., *N*; *N* is the number of important FCs) were applied as explanatory variables in a regression model, and the model was fitted to each fold. **(b)** Feature selection was performed in the inner loop for hyperparameter tuning. With 10-fold CV, the best *k* value was selected based on the prediction performance. The two-variable problem (correlation coefficient and MAE) is solved by identifying the knee point in the Pareto front plot. **(c)** Generated prediction models were applied to the validation dataset. CV: cross-validation, HC: healthy control, SSD: schizophrenia spectrum disorder, FC: functional connectivity.

**Supplementary Figure 3. Probability density curve of voting classifiers**

The abscissa represents the predicted probability of SSD as an output of the classifiers. If a participant’s probability was over 0.5 (vertical dashed line), the participant was classified as SSD, otherwise classified as HC. The ordinate indicates the density at a certain probability. **(a)** Results for all the sites combined in the discovery dataset. **(b)** Results for individual sites in the discovery dataset. COI has only an HC curve since it has no patients with SSD. **(c)** Results for all the sites combined in the validation dataset. **(d)** Results for individual sites in the validation dataset. Since three sites in H (HKH, HRC, and HUH) did not have any patients with SSD, these sites have a curve for HCs only. HC: healthy control, SSD: schizophrenia spectrum disorder, AUC: area under the curve, MCC: Matthews’ correlation coefficient, LASSO: least absolute shrinkage and selection operator, COBRE: Centre of Biomedical Research Excellence.

**Supplementary Figure 4. Probability density curves for HC, SSD, and other disorders reveal classifier specificity for SSD**

**(a)** Results for the LASSO classifiers. **(b)** Results for the voting classifiers. HC: healthy control, SSD: schizophrenia spectrum disorder, MDD: major depressive disorder, ASD: autism spectrum disorder, BP: bipolar disorder, LASSO: least absolute shrinkage and selection operator.
