## Supplementary Tables for "Generalisable functional imaging classifiers of schizophrenia have multifunctionality as trait, state, and staging biomarkers"

**Supplementary Table 1. Participant demographics: individuals with disorders other than SSD**

| **Site** | **Abbr.** | **MDD** | | |  | **ASD** | | |  | **BP** | | |  | **ALL (including HC & SSD)** | | |
| --- | --- | --- | --- | --- | --- | --- | --- | --- | --- | --- | --- | --- | --- | --- | --- | --- |
|  |  | Number | M/F | Age (mean±SD) |  | Number | M/F | Age (mean±SD) |  | Number | M/F | Age (mean±SD) |  | Number | M/F | Age (mean±SD) |
| **Discovery dataset** | | | | | | | | | | | | | | | | |
| Kyoto University (TimTrio) | KUT | 16 | 10/6 | 42.6±12.5 |  | – | – | – |  | – | – | – |  | 300 | 169/131 | 35.5±13.2 |
| Showa University | SWA | – | – | – |  | 115 | 100/15 | 32.1±7.8 |  | – | – | – |  | 235 | 201/34 | 31.4±8.7 |
| Centre of Innovation, Hiroshima University | COI | 70 | 31/39 | 45.0±12.5 |  | – | – | – |  | – | – | – |  | 194 | 77/117 | 49.4±13.5 |
| University of Tokyo | UTO | 62 | 36/26 | 38.7±11.6 |  | 10 | 9/1 | 37.0±9.6 |  | 39 | 24/15 | 34.7±9.1 |  | 316 | 170/146 | 35.7±14.8 |
| Summary |  | 148 | 77/71 | 42.1±12.4 |  | 125 | 109/16 | 32.5±8.0 |  | 39 | 24/15 | 34.7±9.1 |  | 1045 | 617/428 | 37.2±14.3 |
| **Validation dataset** | | | | | | | | | | | | | | | | |
| Kyoto University (Trio) | KTT | – | – | – |  |  | | | | | | |  | See Table 1 | | |
| Kyoto University (Prisma) | KUP | – | – | – |  |  |  |  |  |  |  |  |  | See Table 1 | | |
| Hiroshima University Hospital | HUH | 57 | 32/25 | 43.3±12.2 |  |  |  |  |  |  |  |  |  | 123 | 61/62 | 38.6±13.3 |
| Hiroshima Kajikawa Hospital | HKH | 32 | 19/13 | 44.3±11.3 |  |  |  |  |  |  |  |  |  | 61 | 31/30 | 44.8±10.4 |
| Hiroshima Research Centre | HRC | 16 | 6/10 | 40.5±11.5 |  |  |  |  |  |  |  |  |  | 65 | 19/46 | 41.4±11.5 |
| Centre of Biomedical Research Excellence | COBRE | – | – | – |  |  |  |  |  |  |  |  |  | See Table 1 | | |
| Taipei Medical University | TMU | – | – | – |  |  |  |  |  |  |  |  |  | See Table 1 | | |
| Johns Hopkins University | JHU | – | – | – |  |  |  |  |  |  |  |  |  | See Table 1 | | |
| **Summary** |  | 105 | 57/48 | 43.2±11.8 |  |  |  |  |  |  |  |  |  | 602 | 355/247 | 33.8±11.7 |

HC: healthy control, SSD: schizophrenia spectrum disorder, MDD: major depressive disorder, ASD: autism spectrum disorder, BP: bipolar disorder, Abbr.: abbreviations, SD: standard deviation.

**Supplementary Table 2. Availability of the data on symptom scale scores and medication.**

| **Site** | **Abbr.** | **PDI total score** | | | | | | | |  | **PANSS total score** | |  | **Antipsychotic dose** | |
| --- | --- | --- | --- | --- | --- | --- | --- | --- | --- | --- | --- | --- | --- | --- | --- |
|  |  | HC | |  | SSD | |  | All |  |  | SSD | |  | SSD | |
|  |  | Number | Score  (mean±SD) |  | Number | Score  (mean±SD) |  | Number | Score  (mean±SD) |  | Number | Score  (mean±SD) |  | Number | Dose  (mg, CPZ eq.) (mean±SD) |
| **Discovery dataset** | | | | | | | | | | | | | | | |
| Kyoto University (Tim Trio) | KUT | 105 | 29.7±28.2 |  | 57 | 72.3±50.3 |  | 162 | 44.7±42.6 |  | 60 | 57.2±17.5 |  | 55 | 532.4±384.9 |
| Showa University | SWA | 67 | 23.4±24.1 |  | 18 | 43.1±40.7 |  | 85 | 27.6±29.3 |  | 9 | 63.9±11.2 |  | – | – |
| University of Tokyo | UTO | – | – |  | – | – |  | – | – |  | 36 | 72.3±19.2 |  | – | – |
| **Summary** | | 172 | 27.3±26.8 |  | 75 | 65.3±49.5 |  | 247 | 38.8±39.3 |  | 105 | 62.9±18.9 |  | 55 | 532.4±384.9 |
| **Validation dataset** | | | | | | | | | | | | | | | |
| Kyoto University (Trio) | KTT | 67 | 22.9±21.6 |  | 43 | 107.3±66.5 |  | 110 | 55.9±60.8 |  | 44 | 60.4±20.0 |  | 48 | 610.9±487.8 |
| Kyoto University (Prisma) | KUP | 7 | 31.9±18.7 |  | 8 | 71.8±59.2 |  | 15 | 53.1±48.2 |  | 18 | 62.3±14.0 |  | 16 | 853.9±422.3 |
| Centre of Biomedical Research Excellence | COBRE | – | – |  | – | – |  | – | – |  | 64 | 58.1±13.1 |  | 65 | 350.3±302.4 |
| Johns Hopkins University | JHU | – | – |  | – | – |  | – | – |  | – | – |  | 27 | 430.6±350.0 |
| **Summary** | | 74 | 23.8±21.4 |  | 51 | 101.7±66.1 |  | 125 | 55.6±59.2 |  | 126 | 59.5±15.9 |  | 156 | 496.1±417.9 |

In this table, number of participants with data on PDI total score, PANSS total score, and antipsychotic dose, along with the mean value and standard deviation, are provided. Note that only sites with available data are shown here. Abbr.: abbreviations, HC: healthy control, SSD: schizophrenia spectrum disorder, PDI: Peters Delusion Inventory, PANSS: Positive and Negative Syndrome Scale, SD: standard deviation, CPZ eq.: chlorpromazine equivalent.

**Supplementary Table 3. Imaging parameters of each imaging site.**

|  | **Discovery dataset** | | | |
| --- | --- | --- | --- | --- |
| Site | Kyoto University  (Tim Trio) | Showa University | Centre of Innovation, Hiroshima University | University of Tokyo |
| Abbreviations | KUT | SWA | COI | UTO |
| MRI scanner | Siemens TimTrio | Siemens Verio | Siemens Verio | GE MR750w |
| Magnetic field strength | 3.0 T | | | |
| Channels per coil | 32 | 12 | | 24 |
| Field of view (mm) | 212 × 212 | | | |
| Matrix | 64 × 64 | | | |
| Number of slices | 40 | | | |
| Number of volumes | 240 | | | |
| In-plane resolution (mm) | 3.3125 × 3.3125 | | | |
| Slice thickness (mm) | 3.2 | | | |
| Slice gap (mm) | 0.8 | | | |
| TR (s) | 2.5 | | | |
| TE (ms) | 30 | | | |
| Total scan time | 10'00" | | | |
| Flip angle (degree) | 80 | | | |
| Slice acquisition order | Ascending | | | |
| Phase encoding | P→A | | A→P | P→A |
| Eye condition | Fixated | | | |

|  | **Validation dataset** | | | | | | | |
| --- | --- | --- | --- | --- | --- | --- | --- | --- |
| Site | Kyoto University (Trio) | Kyoto University (Prisma) | Hiroshima University Hospital | Hiroshima Kajikawa Hospital | Hiroshima Research Centre | Centre of Biomedical Research Excellence | Taipei Medical University | Johns Hopkins University |
| Abbreviations | KTT | KUP | HKH | HRC | HUH | COBRE | TMU | JHU |
| MRI scanner | Siemens Trio | Siemens Prisma | Siemens Spectra | GE Signa HDxt | GE Signa HDxt | Siemens TimTrio | GE  MR750w | Philips  Achieva |
| Magnetic field strength | 3.0 T | | | | | | | |
| Channels per coil | 8 | 64 | 12 | 8 | | | 24 | 32 |
| Field of view (mm) | 256 × 192 | 200 × 200 | 192 × 192 | 256 × 256 | | 240 × 240 | 192 × 192 | 240 × 240 |
| Matrix | 64 × 48 | 100 × 100 | 64 × 64 | | | | | 80 × 80 |
| Number of slices | 30 | 72 | 38 | 32 | | 33 | 43 | 36 |
| Number of volumes | 180 | 320 | 107 | 143 | | 150 | 206 | 210 |
| In-plane resolution (mm) | 4 × 4 | 2 × 2 | 3 × 3 | 4 × 4 | | 3.75 × 3.75 | 3 × 3 | 3 × 3 |
| Slice thickness (mm) | 4 | 2 | 3 | 4 | | 3.5 | 3 | 4 |
| Slice gap (mm) | 0 | | | | | 1.05 | 0 | 1 |
| TR (s) | 2 | 0.75 | 2.7 | 2 | | | 2.5 | 2 |
| TE (ms) | 30 | 36.2 | 31 | 27 | | 29 | 30 | 30 |
| Total scan time | 6'00" | 4'10" | 5'00" | 4'46" | 5'00" | 5'00" | 8'35" | 7'00" |
| Flip angle (degree) | 90 | 55 | 90 | | | 75 | 80 | 75 |
| Slice acquisition order | Ascending (interleaved) | | Ascending | Ascending (interleaved) | | | | Ascending |
| Phase encoding | A→P | P→A | A→P | | P→A | A→P | A→P | P→A |
| Eye condition | Fixated | | | | | Not specified | | |

**Supplementary Table 4. Detailed information about 47 important FCs**

| FC No. | ROI 1 | | |  | ROI 2 | | | Selection count | Coefficient for LASSO-LR |
| --- | --- | --- | --- | --- | --- | --- | --- | --- | --- |
|  | Glasser’s label | AAL label | Network |  | Glasser’s label | AAL label | Network |  |  |
| 1 | L.3b | Postcentral_L | Somatomotor |  | L.1 | Postcentral_L | Somatomotor | 17 | −0.045 |
| 2 | L.POS2 | Precuneus_L | Frontoparietal |  | R.SFL | Supp_Motor_Area_R | Language | 20 | −0.070 |
| 3 | L.PCV | Precuneus_L | Posterior-Multimodal |  | L.POS1 | Precuneus_L | Default | 42 | −0.206 |
| 4 | L.POS1 | Precuneus_L | Default |  | L.13l | Frontal_Inf_Orb_L | Frontoparietal | 22 | 0.069 |
| 5 | L.5L | Precuneus_L | Somatomotor |  | R.Cereb | Cerebellum_R | Cerebellar | 19 | 0.086 |
| 6 | L.6ma | Frontal_Sup_L | Cingulo-Opercular |  | R.V8 | Fusiform_R | Visual2 | 21 | 0.102 |
| 7 | L.7Am | Precuneus_L | Cingulo-Opercular |  | R.PFcm | Rolandic_Oper_R | Cingulo-Opercular | 25 | 0.104 |
| 8 | L.1 | Postcentral_L | Somatomotor |  | R.3b | Precentral_R | Somatomotor | 40 | −0.130 |
| 9 | L.2 | Postcentral_L | Somatomotor |  | L.Cereb | Cerebellum_L | Cerebellar | 18 | 0.049 |
| 10 | L.3a | Postcentral_L | Somatomotor |  | L.46 | Frontal_Mid_L | Cingulo-Opercular | 23 | 0.088 |
| 11 | L.6d | Precentral_L | Somatomotor |  | R.Thalamus | Thalamus_R | Subcortical | 18 | 0.089 |
| 12 | L.6mp | Paracentral_Lobule_L | Somatomotor |  | R.Thalamus | Thalamus_R | Subcortical | 24 | 0.124 |
| 13 | L.p32pr | Cingulum_Mid_L | Cingulo-Opercular |  | R.Putamen | Putamen_R | Subcortical | 22 | −0.088 |
| 14 | L.IFSa | Frontal_Inf_Tri_L | Frontoparietal |  | R.AVI | Insula_R | Frontoparietal | 18 | −0.063 |
| 15 | L.11l | Frontal_Sup_Orb_L | Frontoparietal |  | R.V7 | Occipital_Sup_R | Visual2 | 20 | 0.090 |
| 16 | L.MI | Insula_L | Cingulo-Opercular |  | R.2 | Postcentral_R | Somatomotor | 20 | 0.090 |
| 17 | L.MI | Insula_L | Cingulo-Opercular |  | R.Putamen | Putamen_R | Subcortical | 35 | −0.139 |
| 18 | L.FOP1 | Rolandic_Oper_L | Cingulo-Opercular |  | R.Putamen | Putamen_R | Subcortical | 24 | −0.134 |
| 19 | L.PHA1 | Fusiform_L | Default |  | L.Cereb | Cerebellum_L | Cerebellar | 17 | 0.067 |
| 20 | L.STSdp | Temporal_Mid_L | Language |  | L.A4 | Temporal_Sup_L | Auditory | 30 | −0.072 |
| 21 | L.25 | Olfactory_L | Default |  | R.PGi | Angular_R | Default | 21 | 0.065 |
| 22 | L.Ig | Insula_L | Somatomotor |  | L.Cereb | Cerebellum_L | Cerebellar | 33 | 0.123 |
| 23 | L.A4 | Temporal_Sup_L | Auditory |  | R.TPOJ1 | Temporal_Mid_R | Language | 17 | −0.037 |
| 24 | L.A4 | Temporal_Sup_L | Auditory |  | R.Thalamus | Thalamus_R | Subcortical | 45 | 0.184 |
| 25 | R.4 | Precentral_R | Somatomotor |  | R.Thalamus | Thalamus_R | Subcortical | 19 | 0.071 |

(continue to next page)

*(contd.)*

| FC No. | ROI 1 | | |  | ROI 2 | | | Selection count | Coefficient for LASSO-LR |
| --- | --- | --- | --- | --- | --- | --- | --- | --- | --- |
|  | Glasser’s label | AAL label | Network |  | Glasser’s label | AAL label | Network |  |  |
| 26 | R.55b | Precentral_R | Language |  | R.SFL | Supp_Motor_Area_R | Language | 18 | 0.042 |
| 27 | R.RSC | Cingulum_Post_R | Frontoparietal |  | R.SFL | Supp_Motor_Area_R | Language | 18 | −0.083 |
| 28 | R.POS2 | Cuneus_R | Frontoparietal |  | R.SFL | Supp_Motor_Area_R | Language | 20 | −0.077 |
| 29 | R.23d | Cingulum_Mid_R | Default |  | R.FOP3 | Insula_R | Cingulo-Opercular | 20 | −0.088 |
| 30 | R.31pv | Cingulum_Mid_R | Default |  | R.Putamen | Putamen_R | Subcortical | 35 | 0.229 |
| 31 | R.SCEF | Supp_Motor_Area_R | Cingulo-Opercular |  | L.Putamen | Putamen_L | Subcortical | 19 | −0.092 |
| 32 | R.SCEF | Supp_Motor_Area_R | Cingulo-Opercular |  | R.Putamen | Putamen_R | Subcortical | 62 | −0.430 |
| 33 | R.6ma | Frontal_Sup_R | Cingulo-Opercular |  | R.Cereb | Cerebellum_R | Cerebellar | 17 | 0.078 |
| 34 | R.3a | Precentral_R | Somatomotor |  | L.Caudate | Caudate_L | Subcortical | 18 | 0.062 |
| 35 | R.3a | Precentral_R | Somatomotor |  | R.Thalamus | Thalamus_R | Subcortical | 30 | 0.067 |
| 36 | R.6v | Precentral_R | Somatomotor |  | L.Thalamus | Thalamus_L | Subcortical | 27 | 0.088 |
| 37 | R.a24pr | Cingulum_Mid_R | Cingulo-Opercular |  | R.Putamen | Putamen_R | Subcortical | 49 | −0.333 |
| 38 | R.p32pr | Cingulum_Mid_R | Cingulo-Opercular |  | R.Putamen | Putamen_R | Subcortical | 24 | −0.117 |
| 39 | R.10r | Frontal_Med_Orb_R | Default |  | R.PoI1 | Insula_R | Cingulo-Opercular | 17 | −0.066 |
| 40 | R.46 | Frontal_Mid_R | Cingulo-Opercular |  | R.6a | Frontal_Mid_R | Dorsal-attention | 19 | 0.051 |
| 41 | R.6a | Frontal_Mid_R | Dorsal-attention |  | R.PoI1 | Insula_R | Cingulo-Opercular | 36 | 0.133 |
| 42 | R.i6-8 | Frontal_Mid_R | Frontoparietal |  | R.PeEc | ParaHippocampal_R | Ventral-Multimodal | 74 | 0.538 |
| 43 | R.FOP4 | Insula_R | Cingulo-Opercular |  | R.Putamen | Putamen_R | Subcortical | 21 | −0.093 |
| 44 | R.FOP1 | Rolandic_Oper_R | Cingulo-Opercular |  | R.Putamen | Putamen_R | Subcortical | 63 | −0.535 |
| 45 | R.TPOJ1 | Temporal_Mid_R | Language |  | R.Thalamus | Thalamus_R | Subcortical | 22 | 0.057 |
| 46 | R.PFm | Parietal_Inf_R | Frontoparietal |  | R.Ig | Insula_R | Somatomotor | 17 | 0.062 |
| 47 | R.PoI1 | Insula_R | Cingulo-Opercular |  | R.p24 | Cingulum_Ant_R | Cingulo-Opercular | 18 | −0.034 |

This table provides details on each of the 47 important FCs: the constituent ROIs, the selection count (indicating how many times the FC was selected as a feature for prediction by LASSO classifiers), and the mean coefficient value for logistic regression (LR) of LASSO classifiers. FC: functional connectivity, ROI: region of interest, AAL: Anatomical Automatic Labelling, LASSO: least absolute shrinkage and selection operator.

**Supplementary Table 5. Linear regression analysis results for predicting classifier output with potential confounding factors**

|  | **LASSO classifiers** | | |  | **Voting classifiers** | | |
| --- | --- | --- | --- | --- | --- | --- | --- |
|  | *R*^2^ | *F* | *P* |  | *R*^2^ | *F* | *P* |
| Age | 0.013 | 7.79 | **0.0054** |  | 0.013 | 7.39 | **0.0068** |
| Sex | 0.001 | 0.45 | 0.50 |  | 0.000 | 0.24 | 0.62 |
| Framewise displacement | 0.001 | 0.41 | 0.52 |  | 0.005 | 3.12 | 0.078 |
| Head movement parameters | 0.015 | 1.46 | 0.19 |  | 0.015 | 1.44 | 0.20 |
| Antipsychotic dose (SSD only) | 0.024 | 3.40 | 0.067 |  | 0.003 | 0.43 | 0.51 |

We examined on the validation dataset if we could predict the output of the classifiers (probability of SSD) based on the possible confounding factors (in the raw). The linear regression fitting parameters for LASSO and voting classifiers are presented. *P* values that were significant (*P* < 0.05) are highlighted in bold for emphasis. SSD: schizophrenia spectrum disorder, LASSO: least absolute shrinkage and selection operator.

**Supplementary Table 6. Classification performance for different disease severity subgroups**

|  | Disease severity | | | | |  |
| --- | --- | --- | --- | --- | --- | --- |
|  | Mild | Moderate | Marked | Severe | Most severe | **Total** |
| Number | 55 | 36 | 20 | 1 | 0 | 112 |
| LASSO classifier: Sensitivity | 74.5 % | 63.9 % | 95.0 % | 0 % | – | 74.1 % |
| Voting classifier: Sensitivity | 78.2 % | 69.4 % | 95.0 % | 0 % | – | 77.7 % |

Sensitivity to detect patients with SSD is shown for the LASSO and voting classifiers, respectively, assessed across representing varying levels of disease severity. The disease severity was determined based on PANSS total score. PANSS: Positive and Negative Syndrome Scale, LASSO: least absolute shrinkage and selection operator.

**Supplementary Table 7. Analysis of similarity to either HC or SSD based on the classifying results of patients with other disorders .**

|  | LASSO classifiers | |  | Voting classifiers | |
| --- | --- | --- | --- | --- | --- |
|  | *P* value of a two-sided binomial test | Classified as SSD |  | *P* value of a two-sided binomial test | Classified as SSD |
| MDD | **2.24 × 10^-7^** | 33.3 % |  | **4.66 × 10^-3^** | 40.7 % |
| ASD | 0.363 | 45.4 % |  | 1.0 | 50.4 % |
| BP | 0.749 | 53.8 % |  | 0.749 | 53.8 % |

Based on the classifier results of patients with other disorders (MDD, ASD, BP), we assessed whether these patients had high- or low-SSD-like characteristics. Specifically, we conducted a two-sided binomial test on the number of patients classified as SSD. Significance levels (P values) are indicated. Values in bold (*P* < 0.05 denote that the disorder group had significantly high- or low-SSD-ness. The ratio of patients classified as SSD is also provided. HC: healthy control, SSD: schizophrenia spectrum disorder, MDD: major depressive disorder, ASD: autism spectrum disorder, BP: bipolar disorder, LASSO: least absolute shrinkage and selection operator.

**Supplementary Table 8. Comparison of probability distributions between group pairs**

|  | **LASSO classifiers** | | | |  | **Voting classifiers** | | | |
| --- | --- | --- | --- | --- | --- | --- | --- | --- | --- |
|  | HC | MDD | ASD | BP |  | HC | MDD | ASD | BP |
| MDD | **2.18× 10^-14^** |  |  |  |  | **4.80 × 10^-16^** |  |  |  |
| ASD | **2.04 × 10^-7^** | 0.0837 |  |  |  | **3.24 × 10^-9^** | 0.0263 |  |  |
| BP | **1.93 × 10^-6^** | 0.0216 | 0.123 |  |  | **1.01 × 10^-4^** | 0.178 | 0.220 |  |
| SSD | **1.22 × 10^-15^** | **9.99 × 10^-16^** | **2.21 × 10^-11^** | **1.63 × 10^-5^** |  | **1.22 × 10^-15^** | **9.99 × 10^-16^** | **1.39 × 10^-9^** | **0.00169** |

*P* values of the two-sample Kolmogorov-Smirnov test are shown. Statistical significance was assessed with Bonferroni correction, that is, *P* < 0.05/10 = 0.005 was considered significant (emphasised in bold). HC: healthy control, SSD: schizophrenia spectrum disorder, MDD: major depressive disorder, ASD: autism spectrum disorder, BP: bipolar disorder, LASSO: least absolute shrinkage and selection operator.

**Supplementary Table 9. Individual FCs significantly associated with PANSS factorial scores and regression parameters**

A. Min-max transformation (three factor model)

| FC | Diagnosis | | |  | PANSS positive | | |  | PANSS negative | | |
| --- | --- | --- | --- | --- | --- | --- | --- | --- | --- | --- | --- |
|  | coefficient | *t* | *P* |  | coefficient | *t* | *P* |  | coefficient | *t* | *P* |

| #41 | 0.060 | 1.83 | 0.067 |  | **0.161** | **2.19** | **0.029** |  | − 0.048 | −0.67 | 0.50 |
| --- | --- | --- | --- | --- | --- | --- | --- | --- | --- | --- | --- |
| #45 | **0.135** | **3.68** | **2.5×10**^−4^ |  | **0.191** | **2.32** | **0.021** |  | − 0.141 | −1.79 | 0.074 |
| #1 | −0.090 | −1.62 | 0.11 |  | 0.017 | 0.14 | 0.89 |  | − **0.244** | −**2.05** | **0.040** |
| #8 | −0.092 | −1.77 | 0.078 |  | 0.161 | 1.38 | 0.17 |  | − **0.291** | −**2.60** | **0.0094** |

B. Binarisation (three factor model)

| FC | Diagnosis | | |  | PANSS positive | | |  | PANSS negative | | |
| --- | --- | --- | --- | --- | --- | --- | --- | --- | --- | --- | --- |
|  | coefficient | *t* | *P* |  | coefficient | *t* | *P* |  | coefficient | *t* | *P* |

| #33 | 0.002 | 0.05 | 0.96 |  | **0.114** | **2.80** | **0.0052** |  | −0.045 | −1.10 | 0.27 |
| --- | --- | --- | --- | --- | --- | --- | --- | --- | --- | --- | --- |
| #45 | **0.155** | **5.23** | **2.0× 10**^−7^ |  | **0.080** | **2.08** | **0.038** |  | −0.073 | −1.89 | 0.060 |
| #2 | −0.001 | −0.05 | 0.96 |  | −0.001 | −0.02 | 0.99 |  | −**0.094** | −**2.83** | **0.0047** |
| #8 | **−0.100** | **−2.37** | **0.018** |  | 0.023 | 0.41 | 0.68 |  | −**0.108** | −1.96 | **0.0497** |
| #28 | −0.012 | −0.45 | 0.66 |  | −0.009 | −0.26 | 0.79 |  | **−0.113** | **−3.36** | **8.2×10^−4^** |

C. Min-max transformation (five factor model)

| FC | Diagnosis | | |  | Positive factor | | |  | Negative factor | | |  | Disorganised factor | | |  | Excited factor | | |  | Depressed factor | | |
| --- | --- | --- | --- | --- | --- | --- | --- | --- | --- | --- | --- | --- | --- | --- | --- | --- | --- | --- | --- | --- | --- | --- | --- |
|  | coeff. | *t* | *P* |  | coeff. | *t* | *P* |  | coeff. | *t* | *P* |  | coeff. | *t* | *P* |  | coeff. | *t* | *P* |  | coeff. | *t* | *P* |
| #5 | 0.074 | 1.60 | 0.11 |  | **0.267** | **2.05** | **0.041** |  | 0.030 | 0.09 | 0.74 |  | -0.091 | -0.68 | 0.498 |  | 0.013 | 0.12 | 0.91 |  | -0.283 | -2.11 | 0.035 |
| #42 | 0.067 | 1.89 | 0.060 |  | **0.304** | **3.04** | **0.0024** |  | -0.045 | -0.64 | 0.52 |  | -0.167 | -1.62 | 0.11 |  | -0.040 | -0.47 | 0.64 |  | -0.066 | -0.65 | 0.52 |
| #45 | **0.157** | **3.60** | **3.4×10^-4^** |  | **0.248** | **2.00** | **0.045** |  | -0.093 | -1.07 | 0.28 |  | -0.010 | -0.08 | 0.94 |  | -0.043 | -0.41 | 0.69 |  | -0.074 | -0.58 | 0.56 |
| #17 | **-0.202** | **-5.55** | **3.5×10^-8^** |  | -0.033 | -0.32 | 0.75 |  | 0.041 | 0.57 | 0.57 |  | 0.157 | 1.48 | 0.14 |  | **-0.176** | **-2.01** | **0.045** |  | 0.082 | 0.78 | 0.44 |
| #18 | **-0.116** | **-3.12** | **0.0016** |  | 0.035 | 0.34 | 0.73 |  | -0.045 | -0.63 | 0.53 |  | 0.054 | 0.51 | 0.61 |  | **-0.181** | **-2.05** | **0.040** |  | 0.153 | 1.44 | 0.15 |
| #41 | 0.070 | 1.79 | 0.074 |  | 0.121 | 1.09 | 0.27 |  | -0.075 | -0.97 | 0.33 |  | -0.066 | -0.58 | 0.56 |  | **0.215** | **2.29** | **0.022** |  | -0.035 | -0.031 | 0.76 |
| #43 | **-0.243** | **-6.78** | **1.9×10^-11^** |  | 0.143 | 1.41 | 0.16 |  | 0.092 | 1.30 | 0.19 |  | 0.186 | 1.79 | 0.075 |  | **-0.258** | **-2.99** | **0.0029** |  | 0.011 | 0.10 | 0.92 |
| #27 | -0.037 | -0.91 | 0.36 |  | 0.080 | 0.70 | 0.49 |  | -0.046 | -0.57 | 0.57 |  | -0.028 | -0.24 | 0.81 |  | 0.188 | 1.92 | 0.056 |  | **-0.351** | **-2.96** | **0.0031** |
| #28 | -0.009 | -0.23 | 0.82 |  | -0.021 | -0.19 | 0.85 |  | -0.14 | -1.85 | 0.065 |  | -0.008 | -0.07 | 0.95 |  | 0.169 | 1.84 | 0.067 |  | **-0.219** | **-1.97** | **0.0497** |

D. Binarisation (five factor model)

| FC | Diagnosis | | |  | Positive factor | | |  | Negative factor | | |  | Disorganised factor | | |  | Excited factor | | |  | Depressed factor | | |
| --- | --- | --- | --- | --- | --- | --- | --- | --- | --- | --- | --- | --- | --- | --- | --- | --- | --- | --- | --- | --- | --- | --- | --- |
|  | coeff. | *t* | *P* |  | coeff. | *t* | *P* |  | coeff. | *t* | *P* |  | coeff. | *t* | *P* |  | coeff. | *t* | *P* |  | coeff. | *t* | *P* |
| #3 | -0.014 | -0.35 | 0.73 |  | **-0.119** | **-2.32** | **0.020** |  | 0.036 | 0.76 | 0.45 |  | 0.024 | 0.48 | 0.63 |  | 0.074 | 0.89 | 0.37 |  | -0.010 | -0.19 | 0.85 |
| #8 | -0.084 | -1.64 | 0.10 |  | 0.070 | 1.06 | 0.29 |  | **-0.127** | **-2.07** | **0.039** |  | **-0.127** | **-2.03** | **0.043** |  | 0.088 | 0.82 | 0.41 |  | 0.096 | 1.42 | 0.16 |
| #2 | 0.012 | 0.373 | 0.71 |  | -0.036 | -0.89 | 0.37 |  | -0.082 | -2.20 | 0.028 |  | -0.010 | -0.25 | 0.80 |  | **0.168** | **2.57** | **0.010** |  | -0.023 | 0.55 | 0.58 |
| #14 | **-0.095** | **-3.21** | **0.0014** |  | 0.078 | 2.02 | 0.044 |  | 0.050 | 1.40 | 0.16 |  | 0.061 | 1.68 | 0.094 |  | **-0.196** | **-3.14** | **0.0017** |  | -0.013 | -0.32 | 0.75 |
| #18 | **-0.119** | **-3.96** | **8.1×10^-5^** |  | -0.046 | -1.18 | 0.24 |  | -0.029 | -0.81 | 0.42 |  | 0.083 | 2.23 | 0.026 |  | **-0.186** | **-2.93** | **0.0034** |  | 0.076 | 1.90 | 0.057 |
| #38 | **-0.172** | **-5.66** | **2.0×10^-8^** |  | -0.024 | -0.61 | 0.54 |  | -0.015 | 0.40 | 0.69 |  | 0.054 | 1.45 | 0.15 |  | **-0.132** | **-2.05** | **0.040** |  | 0.002 | 0.05 | 0.96 |
| #41 | **0.070** | **2.18** | **0.030** |  | -0.034 | -0.80 | 0.42 |  | -0.034 | -0.87 | 0.38 |  | 0.015 | 0.38 | 0.70 |  | **0.180** | **2.65** | **0.0082** |  | 0.067 | 1.57 | 0.12 |
| #43 | **-0.203** | **-6.84** | **1.3×10^-11^** |  | 0.012 | 0.30 | 0.76 |  | 0.046 | 1.29 | 0.20 |  | 0.067 | 1.84 | 0.066 |  | **-0.173** | **-2.78** | **0.0055** |  | -0.009 | -0.22 | 0.83 |
| #44 | **-0.182** | **-5.88** | **5.2×10^-9^** |  | 0.011 | 0.27 | 0.79 |  | 0.061 | 1.64 | 0.10 |  | 0.080 | 2.10 | 0.036 |  | **-0.197** | **-3.02** | **0.0026** |  | -0.009 | -0.22 | 0.83 |
| #27 | -0.031 | -0.93 | 0.35 |  | 0.016 | 0.36 | 0.72 |  | -0.021 | -0.53 | 0.60 |  | -0.048 | -1.15 | 0.25 |  | 0.282 | 4.00 | 6.9×10^-5^ |  | **-0.121** | **-2.71** | **0.0068** |

Results of multiple regression analyses to identify FCs associated with PANSS factors. With each factorial model and each conversion method, significantly associated FCs were presented. Coefficients, *t*-values, and *P* values for explanatory variables that were significant (*P* < 0.05) are highlighted in bold letters. Note that explanatory variables for PANSS factors were considered significant only when the signs of their coefficients were the same as that of the diagnosis. FC: functional connectivity, coeff.: coefficient, PANSS: Positive and Negative Syndrome Scale.

**Supplementary Table 10. The AAL ROIs frequently included in the 47 important FCs.**

| **ROI (AAL)** | **count** | | |  | **ROI (AAL)** | **count** | | |
| --- | --- | --- | --- | --- | --- | --- | --- | --- |
|  | **total** | **left** | **right** |  |  | **total** | **left** | **right** |
| Insula | 10 | 3 | 7 |  | Caudate | 1 | 1 | 0 |
| Putamen | 10 | 1 | 9 |  | Cingulum_Ant | 1 | 0 | 1 |
| Precentral | 7 | 1 | 6 |  | Cingulum_Post | 1 | 0 | 1 |
| Thalamus | 7 | 1 | 6 |  | Cuneus | 1 | 0 | 1 |
| Postcentral | 6 | 5 | 1 |  | Frontal_Inf_Orb | 1 | 1 | 0 |
| Precuneus | 6 | 6 | 0 |  | Frontal_Inf_Tri | 1 | 1 | 0 |
| Supp_Motor_Area | 6 | 0 | 6 |  | Frontal_Med_Orb | 1 | 0 | 1 |
| Cerebellum | 5 | 3 | 2 |  | Frontal_Sup | 1 | 1 | 0 |
| Cingulum_Mid | 5 | 1 | 4 |  | Frontal_Sup_Orb | 1 | 1 | 0 |
| Frontal_Mid | 5 | 1 | 4 |  | Frontal_Sup | 1 | 0 | 1 |
| Rolandic_Oper | 3 | 1 | 2 |  | Occipital_Sup | 1 | 0 | 1 |
| Temporal_Mid | 3 | 1 | 2 |  | Olfactory | 1 | 1 | 0 |
| Temporal_Sup | 3 | 3 | 0 |  | Paracentral_Lobule | 1 | 1 | 0 |
| Fusiform | 2 | 1 | 1 |  | ParaHippocampal | 1 | 0 | 1 |
| Angular | 1 | 0 | 1 |  | Parietal_Inf | 1 | 0 | 1 |

Within the important FCs delineated in **Supplementary Table 2**, the count of FCs including a specific AAL ROI is presented in descending order. ROI: region of interest, AAL: Anatomical Automatic Labelling, FC: functional connectivity.
