## Supplementary Texts for "Generalisable functional imaging classifiers of schizophrenia have multifunctionality as trait, state, and staging biomarkers"

**Supplementary text 1: Travelling subject harmonisation method**

We estimated the participant factor (***p***), measurement bias (***m***), sampling biases (***s***_hc_, ***s***_ssd_, ***s***_mdd_), and psychiatric disorder factor (***d***) by fitting the regression model to the FC values of all participants from both the discovery and travelling subject datasets. As patients with autism spectrum disorder (ASD) or bipolar disorder (BP) were from only one site, respectively, we did not need to assume a measurement bias for ASD and BP. For each FC, the regression model is formulated as follows:

$Connectivity={x_{m}}^{T}m+{x_{s_{hc}}}^{T}s_{hc}+{x_{s_{ssd}}}^{T}s_{ssd}+{{{x_{s_{mdd}}}^{T}s_{mdd}+x}_{d}}^{T}d+{x_{p}}^{T}p+const+e$ (Eq. S1)

$$such that\sum_{j}^{9} p_{j}=0,\sum_{k}^{4} m_{k}=0,\sum_{k}^{4} {s_{hc}}_{k}=0,\sum_{k}^{3} {s_{ssd}}_{k}=0,{\sum_{k}^{3} {s_{mdd}}_{k}=0,d}_{1}\left( HC \right)=0$$

where $m$ is the measurement bias (4 sites × 1), $s_{hc}$ is the sampling bias of HCs (4 sites × 1), $s_{ssd}$ is the sampling bias of patients with SSD, $s_{mdd}$ is the sampling bias of patients with major depressive disorder (MDD), $d$ is the disorder factor (2 × 1), $p$ is the participant factor (9 travelling subjects × 1), $const$ is the average FC value across all participants from all the sites, and $e \sim\mathcal{N}\left( 0,\gamma^{-1} \right)$ denotes noise. A harmonised FC value was obtained by subtracting the estimated measurement bias from the following equation:

${Connectivity}^{Harmonised}=Connectivity-{x_{m}}^{T}\grave{m}$	(Eq. S2)

where $\grave{m}$ denotes the estimated measurement bias.

**Supplementary text 2: Logistic regression analysis with LASSO and hyper-parameter tuning**

The following is a logistic function for estimating the probability of classifying a participant in the SSD class:

$P_{sub}\left( y_{sub}=1 | c_{sub};w \right)=\frac{1}{1+exp\left( -w^{T}c_{sub} \right)}$	(Eq. S3)

where $y_{sub}$ is the class label (SSD, *y* = 1; HC, *y* = 0) of a participant, $c_{sub}$ is the FC vector for a given participant, and $w$ is a weight vector. The weight $w$ was determined such that $J\left( w \right)$ in the following formulae was minimised:

$J\left( w \right)=\frac{-1}{n_{sub}}\sum_{j=1}^{n_{sub}} logP_{j}\left( y_{j}=1 | c_{j};w \right)+\lambda\left\| w \right\|_{1}$ (Eq. S4)

$\left\| w \right\|_{1}=\sum_{i}^{N} \left| w_{i} \right|$ (Eq. S5)

where $\lambda$ is a hyperparameter that controls the amount of shrinkage applied to the estimate. To estimate weights of the logistic regression and the regularisation hyperparameter $\lambda$, we employed a nested 10-fold cross-validation (CV) scheme (**Supplementary Fig. 1**). To determine $\lambda$ in the inner loop, we executed *lassoglm* function in MATLAB (R2019a, Mathworks, USA) by setting “NumLambda” to 25 and “CV” to 10. The value just large enough that the only optimal solution is the all-zeroes vector was defined as $\lambda_{max}$. From 25 values of $\lambda$, which were placed at equal intervals between 0 and $\lambda_{max}$, the optimal $\lambda$ was selected according to the one-standard-error rule (selecting the largest value within the standard deviation of the minimum prediction error).

**Supplementary text 3: Matthews correlation coefficient (MCC)**

MCC is one of the most balanced and informative metrics that can measure the prediction performance of binary classification as a single evaluation score^1,2^. This method is particularly accurate for data with classes of very different sizes. It is calculated using the following formula:

$$MCC=\frac{TP\cdot TN-FP\cdot FN}{\sqrt{(TP+FP)(TP+FN)(TN+FP)(TN+FN)}}$$

where *TP* is a true positive, *TN* is a true negative, *FP* is a false positive, and *FN* is a false negative in a confusion matrix.

It returns a value between -1 and 1, with 1 being a perfect prediction, 0 indicating no better than random prediction, and -1 representing total disagreement between prediction and observation.

**Supplementary text 4 : Detailed procedure to build voting classifiers**

To build voting classifiers, we incorporated support vector machine (SVM)^3^, random forest (RF)^4^, light gradient boosting machine (LGBM)^5^, and multi-layer perceptron (MLP)^6^ as representative algorithms. The machine-learning packages for each algorithm were LightGBM version 3.2.1 for LGBM, Keras version 2.6.0 for MLP, and scikit-learn version 0.24.1 for SVM and RF. For each algorithm, we conducted CV, subsampling with undersampling, and training on the discovery dataset following the same procedure used to build the LASSO classifiers. Hyperparameters were tuned using Optuna version 2.9.1 in 10-fold CV within the inner loop of the nested CV. We constructed 10 classifiers by repeating the subsampling and training of the model 10 times. In each fold of the outer-loop CV, we predicted the class of each participant in a test set using these 10 classifiers (for LASSO, LGBM, and MLP, the default output was in the form of a probability value. Therefore, when the probability value was > 0.5, the predicted class was ‘SSD’ and vice versa). Similar to voting, we counted the number of times the participant was predicted to have an SSD by ten classifiers for each of the five machine learning algorithms (maximal count: 10 classifiers × 5 algorithms = 50). When the count exceeded 25, the participant was considered to have SSD. Using 10-time subsampling and 10-fold CV for each of the five algorithms, we obtained 500 classifiers. Next, we predicted the diagnostic class in the validation dataset using each of these classifiers, and the final prediction by voting was determined as the class predicted by the majority of the classifiers (> 250).

**Supplementary text 5: Feature selection procedure for building prediction models of clinical scale score**

We sought the most suitable number of features (important FCs) for the hyperparameter-tuning step. During this feature selection process, we initially prioritised the *N* important FCs primarily by the selection count of LASSO classifiers (the number of LASSO classifiers that selected a specific FC as an explanatory variable in their models, on a scale of 0-100) primarily, and secondarily by the absolute value of the classifier weight (the mean coefficient of logistic regression with LASSO in SSD classifiers).

After selecting the top *k* FCs (where *k* = 1, 2, ..., *N*) from the ordered list, we used them as features for the regression models and evaluated the model predictability in the inner loop of the 10-fold CV. The evaluation metrics included Pearson’s correlation coefficient (*r*) and mean absolute error (MAE). We aimed to determine the most suitable *k* that achieved both adequately high *r* and low MAE simultaneously.

To address this two-variable problem, we identified the Pareto frontier^7^ of the plots for each *k* based on the *r* and MAE values. We then selected the knee point of the Pareto frontier as the ideal combination of *r* and MAE. In cases where the Pareto frontier had three or more plots, we attempted to identify a knee point using the Python module *kneed* version 0.7.0^8^. The default sensitivity settings were set to 1. If a knee point was not identified, the sensitivity was halved and repeated until the knee point was identified. When a Pareto frontier had only two plots, the plot with fewer FCs was considered the best solution. In cases where a Pareto frontier had only one plot, the plot was deemed the solution.

Ultimately, the *k* corresponding to the knee point was determined to be the most suitable number for folding.

**Supplementary text 6: Assessment of confounding effects on classifiers’ performance**

We also considered the possibility that the performance of the classifiers may be influenced by confounding factors. To assess this, we examined whether we could predict the output of the classifiers (probability of schizophrenia spectrum disorder (SSD)) based on sex, frame-wise displacement (FD), or a combination of head movement parameters during MRI scanning (mean values of transverse movements in *x*, *y*, *z* directions, yaw, pitch, and roll) through linear regression on the validation dataset.

The classifier’s outputs could not be predicted with sex, FD, or head movement parameters but could be predicted with age (**Supplementary Table 5**). However, this may be attributed to group differences in age. Therefore, we further assessed whether a comparable generalisation performance could be achieved in age-matched samples. We prepared 100 age-matched subsamples from a validation dataset. As a result, for age-matched subsamples, the performance of classifiers based on least absolute shrinkage and selection operator (LASSO) was similar to that for the whole validation dataset, both in area under the receiver operating characteristic curve (AUC) (0.825 ± 0.012 [mean ± standard deviation (SD)]) and in Matthews’ correlation coefficient (MCC) (0.506 ± 0.023 [mean ± SD]), while the voting classifiers performed similarly in AUC (0.842 ± 0.011) but slightly higher in MCC (0.537 ± 0.022) compared to those for the whole validation dataset.

Additionally, we tested whether the classifier output could be predicted using the antipsychotic dose in patients with SSD. Consequently, we could not make predictions for either the LASSO or the voting classifiers.
